# Suvorexant Inversely Affects Phosphorylated Tau in Human Cerebrospinal Fluid and Plasma

**DOI:** 10.64898/2026.11.03.26367196

**Authors:** Taylor J. Pedersen, Jinsheng Yu, Alan Li, Laura Ibanez, Omonigho M. Bubu, Brendan P. Lucey

## Abstract

Alzheimer disease (AD) is characterized by amyloid-β plaques and neurofibrillary tangles of tau. Suvorexant, a dual orexin receptor antagonist, was previously shown to acutely reduce the ratio of phosphorylated tau-181 to unphosphorylated tau-181 (pT181/T181) in cerebrospinal fluid (CSF). However, its effects on plasma pT181/T181 and broader proteomic networks remain unknown. Here, we conducted a secondary analysis of samples from a randomized, placebo-controlled trial of 25 participants receiving placebo or suvorexant 20 mg. Paired CSF and plasma samples were collected longitudinally via indwelling intrathecal and intravenous catheters. Plasma AD biomarkers were quantified by mass spectrometry, and proteomic profiling was performed using the SomaScan 11K assay on samples from hours 0, 16, and 24. Suvorexant increased pT181/T181 in plasma while decreasing it in CSF between 08:00–16:00. At the pathway level, suvorexant increased the extracellular abundance of proteins associated with translation, RNA binding, cytoskeletal organization and endosomal sorting, centrally at 16 hours and peripherally at 24 hours, and decreased cytokine and receptor-ligand signaling and serine protease activity in plasma at 24 hours. As plasma biomarkers are increasingly used for clinical screening and trial enrollment, factors that influence their levels, including medications, must be identified and their effects on interpretation understood.

## INTRODUCTION

Alzheimer disease (AD) is a progressive neurodegenerative disorder marked by the accumulation of extracellular amyloid-β (Aβ) plaques and intraneuronal tau tangles, leading to synaptic dysfunction, cognitive decline, and eventually death.^1^ Recent advances in biofluid biomarker development have enabled increasingly sensitive and specific detection of AD pathology using blood-based markers, complementing well-established cerebrospinal fluid (CSF) and neuroimaging modalities.^2, 3^ CSF biomarkers such as Aβ42, phosphorylated tau (p-tau) and total tau (t-tau) reflect key aspects of AD pathophysiology and are widely used in both clinical and research settings. These markers, however, require lumbar puncture and are not always feasible for large-scale or longitudinal studies. In contrast, plasma-based biomarkers, including Aβ42 and p-tau forms, are minimally invasive and reach a level of analytical performance that allows for reliable detection of early pathological changes in AD.^4, 5^ However, questions remain about the concordance and timing of changes in AD biomarkers between plasma and CSF, particularly in response to pharmacological interventions.

Growing evidence suggests the orexinergic system is a critical regulator in modulating Aβ and tau dynamics.^6, 7^ Orexins (also known as hypocretins) are hypothalamic neuropeptides that promote arousal and regulate sleep-wake transitions.^8^ In animal models, orexin antagonism reduces Aβ levels and plaque formation while orexin administration increases interstitial Aβ levels.^9^ Furthermore, blocking orexin also decreases tau-mediated neurodegeneration through reduced phosphorylation of tau.^10^ Suvorexant, a dual orexin receptor antagonist (DORA), is approved by the Food and Drug Administration (FDA) for the treatment of insomnia.^11^ A recent randomized controlled trial in cognitively unimpaired adults demonstrated that a single dose of suvorexant 20 mg acutely reduced CSF Aβ levels approximately 10-20% and decreased the ratio of phosphorylated tau-threonine-181 to unphosphorylated tau-threonine-181 (pT181/T181), a site linked to early AD pathology, approximately 10-15% compared to placebo.^12^ These changes occurred in the absence of measurable differences in total sleep time or sleep architecture, suggesting a direct pharmacodynamic effect of orexin receptor antagonism on CSF AD biomarkers. While these results provide compelling evidence that suvorexant modulates key AD biomarkers in the central nervous system (CNS), a number of critical questions remain such as suvorexant’s effect on plasma AD biomarkers, the degree to which the CNS effects translate to peripheral compartments, and the molecular breadth of suvorexant’s effects. In addition to understanding the effects of suvorexant on AD biomarkers, understanding the systems-level proteomic effects could reveal mechanistic insights into orexin modulation and inform future prevention strategies.

High-throughput proteomic technologies offer a powerful approach to characterize the broad biological effects of CNS-active agents such as suvorexant. SomaScan, developed by SomaLogic, uses modified DNA aptamers (SOMAmers) to quantify 11,000 aptamers with high specificity and sensitivity from small-volume plasma or CSF samples. It covers a broad range of biological processes including inflammation, synaptic function, metabolism, vascular health, and phosphorylation cascades. Complementary to this, NUcleic acid Linked Immuno-Sandwich Assay (NULISA) is a bead-based, high-sensitivity immunoassay platform optimized for low-abundance protein detection. Together, these platforms enable accurate measurement of core AD biomarkers, deep profiling of known AD-relevant pathways, and the discovery of novel protein networks altered by suvorexant in both CSF and plasma.

This study tests the acute effect of suvorexant 20 mg on pT181/T181 and other AD biomarkers in paired CSF and plasma samples compared to placebo. The results will establish the degree to which peripheral changes in AD biomarkers reflect central dynamics, a critical issue for blood-based diagnostic development and drug monitoring. Then, we will determine whether suvorexant 20 mg alters individual protein levels and proteomic networks in plasma and CSF relative to placebo. We aim to identify proteomic networks whose response to suvorexant may be compartment-specific or correlated with changes in p-tau levels. By integrating targeted biomarkers and untargeted systems proteomics, this study aims to develop a comprehensive molecular map of suvorexant’s acute effects on the human proteome in both central and peripheral compartments. Findings from this work will inform both the utility of suvorexant as a repurposed disease-modifying therapeutic and the development of fluid biomarkers for early pharmacodynamic effects in prevention trials.

## RESULTS

### Differential Effects of Suvorexant on CSF and Plasma pT181/T181

Suvorexant 20 mg was previously reported to decrease pT181/T181 in CSF compared to placebo.^12^ In plasma, pT181/T181 initially decreased in both placebo and suvorexant groups before rebounding after hour 6 in the suvorexant treated participants (Figure 1). The greatest group difference in plasma pT181/T181 occurred approximately 15 hours post-dose (12:00), approximately when the greatest group difference was seen in CSF. Changes in CSF and plasma were seen with the pT181/T181 ratio rather than individual pT181 or T181. There were no group differences in CSF or plasma T181, strongly supporting that suvorexant is associated with decreased tau phosphorylation occupancy at threonine-181 in CSF and increased occupancy in plasma. Further, there were no group differences in plasma unphosphorylated tau at threonine-217 (T217) which showed similar change as plasma T181 (Supplementary Figure 1). Plotting the area under the curve (AUC) of CSF and plasma pT181/T181 percent change from hour 0 between hours 12-20 (08:00-16:00) demonstrated a significant inverse correlation (r = -0.42, p = 0.039; Figure 2) indicating that as pT181/T181 decreased in CSF, the ratio increased in plasma. Unlike in CSF, plasma Aβ40 and Aβ42 levels did not change significantly after treatment with suvorexant.

**Figure 1.**
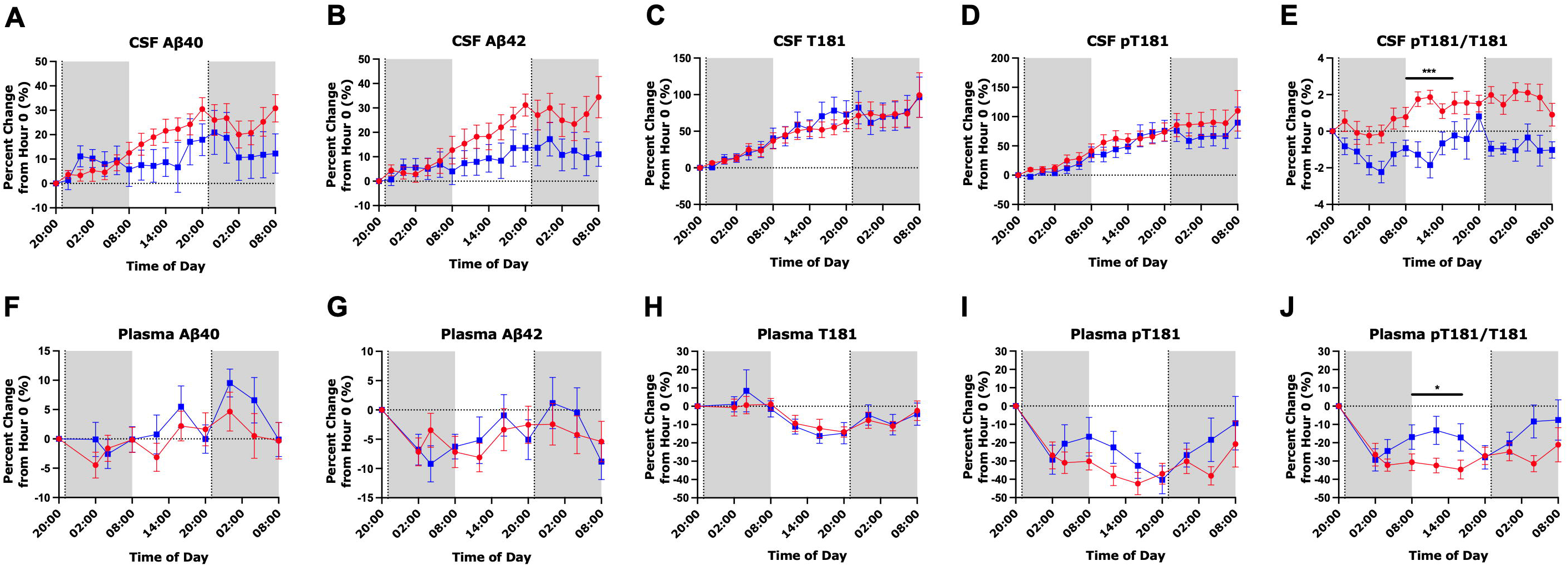
CSF and plasma Alzheimer disease biomarker changes in cognitively unimpaired participants treated with suvorexant 20 mg (blue squares) compared to placebo (red circles) over two consecutive nights. Percent change from hour 0 (20:00, day 1) in CSF Aβ40 (A), CSF Aβ42 (B), CSF unphosphorylated tau-181 (T181) (C), CSF phosphorylated tau-181 (pT181) (D), CSF pT181/T181 ratio (E), plasma Aβ40 (F), plasma Aβ42 (G), plasma T181 (H), plasma pT181 (I), and plasma pT181/T181 ratio (J). Placebo N = 13 except for plasma tau and p-tau measures which were N=12; suvorexant N =12. Points are group mean ± SEM. Shaded regions indicate the scheduled dark period (20:00–08:00); vertical dotted lines indicate administration of placebo or suvorexant at hour 1 (21:00, day 1) and hour 25 (21:00, day 2). CSF data are shown for comparison and were previously reported.^12^ ***p < 0.001; *p < 0.05

**Figure 2.**
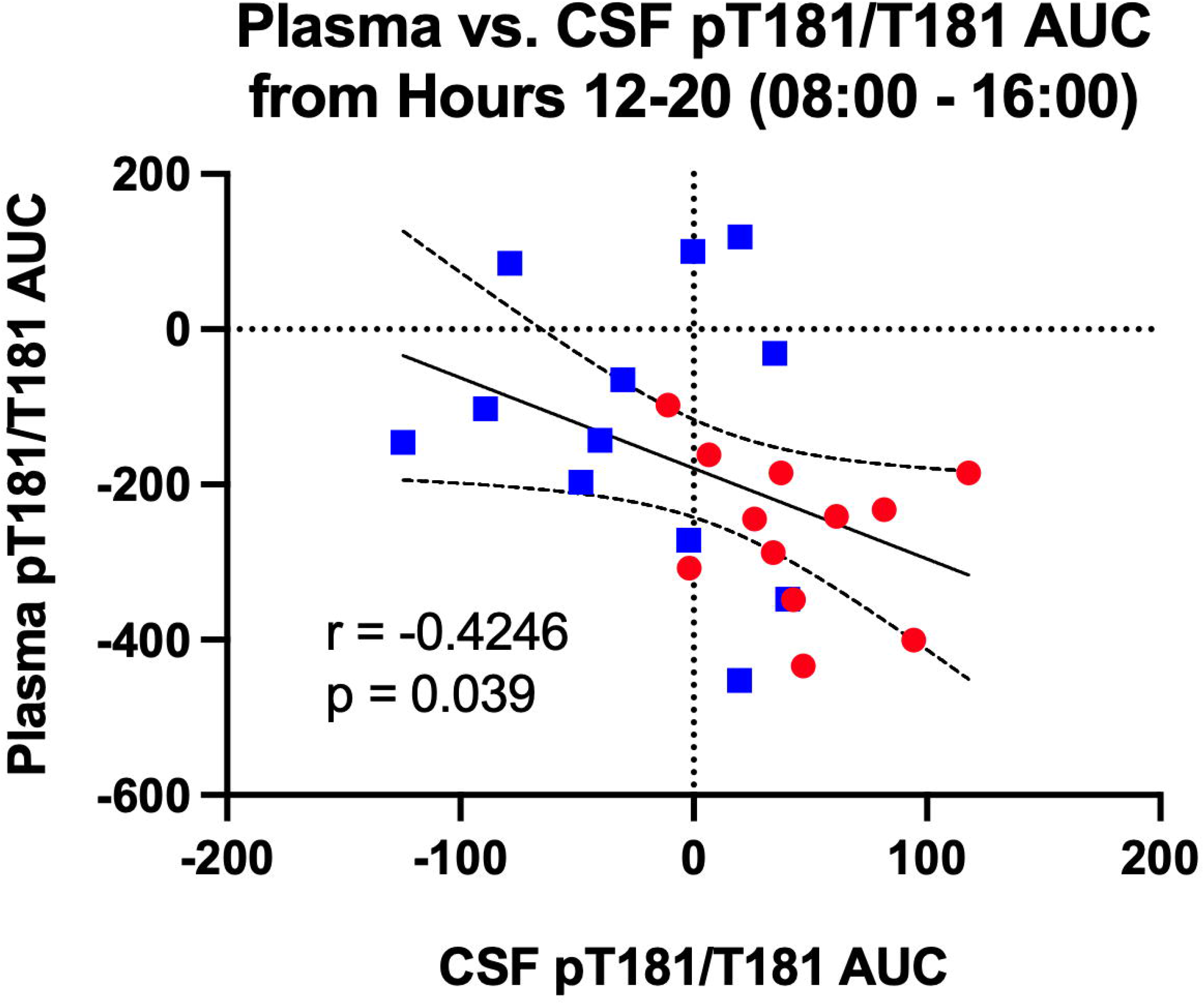
Individual CSF and plasma pT181/T181 responses to suvorexant are inversely correlated. Area under the curve (AUC) of plasma pT181/T181 plotted against AUC of CSF pT181/T181, each calculated from the percent-change-from-hour-0 trajectory over hours 12–20 (08:00–16:00), the interval of greatest group separation. Each symbol is one participant (placebo, red circles, N = 12; suvorexant 20 mg, blue squares, N = 12). Solid line, least-squares linear regression fit to all 24 participants; dashed lines, 95% confidence band. Participants with a larger decrease in CSF pT181/T181 showed a larger increase in plasma pT181/T181 (r = −0.4246, p = 0.039).

The temporal pattern of biomarker changes between biofluid compartments suggested at least three independent hypotheses: 1) suvorexant increases clearance of pT181 from the central to the peripheral compartment; 2) suvorexant exerts different effects on tau phosphorylation in CSF and plasma; and 3) suvorexant alters peripheral processing of tau such that phosphorylated and unphosphorylated T181 are differentially changed in plasma. To investigate these hypotheses and other potential mechanisms, proteomics was measured using SomaScan and NULISA. Although individual protein levels changed at both time points in CSF and plasma, none of these remained significant after multiple test correction using the Benjamini-Hochberg false discovery rate (FDR) procedure (Figure 3; Supplementary Figures 2 and 3). There were several individual proteins with similar changes by both assays at hour 24 in CSF (C-C motif chemokine ligand 2 (CCL2), interleukin-6 (IL-6), interleukin-8 (IL-8), C-X-C motif chemokine ligand 1 (CXCL1)) and plasma (malate dehydrogenase 1, peroxiredoxin 6, *a*-synuclein (SNCA) (total), *a*-synuclein oligo-SNCA (oligomeric)). C-C motif chemokine ligand 13 (CCL13), however, decreased in CSF at hour 24 when measured by SomaScan and increased when measured by NULISA.

**Figure 3.**
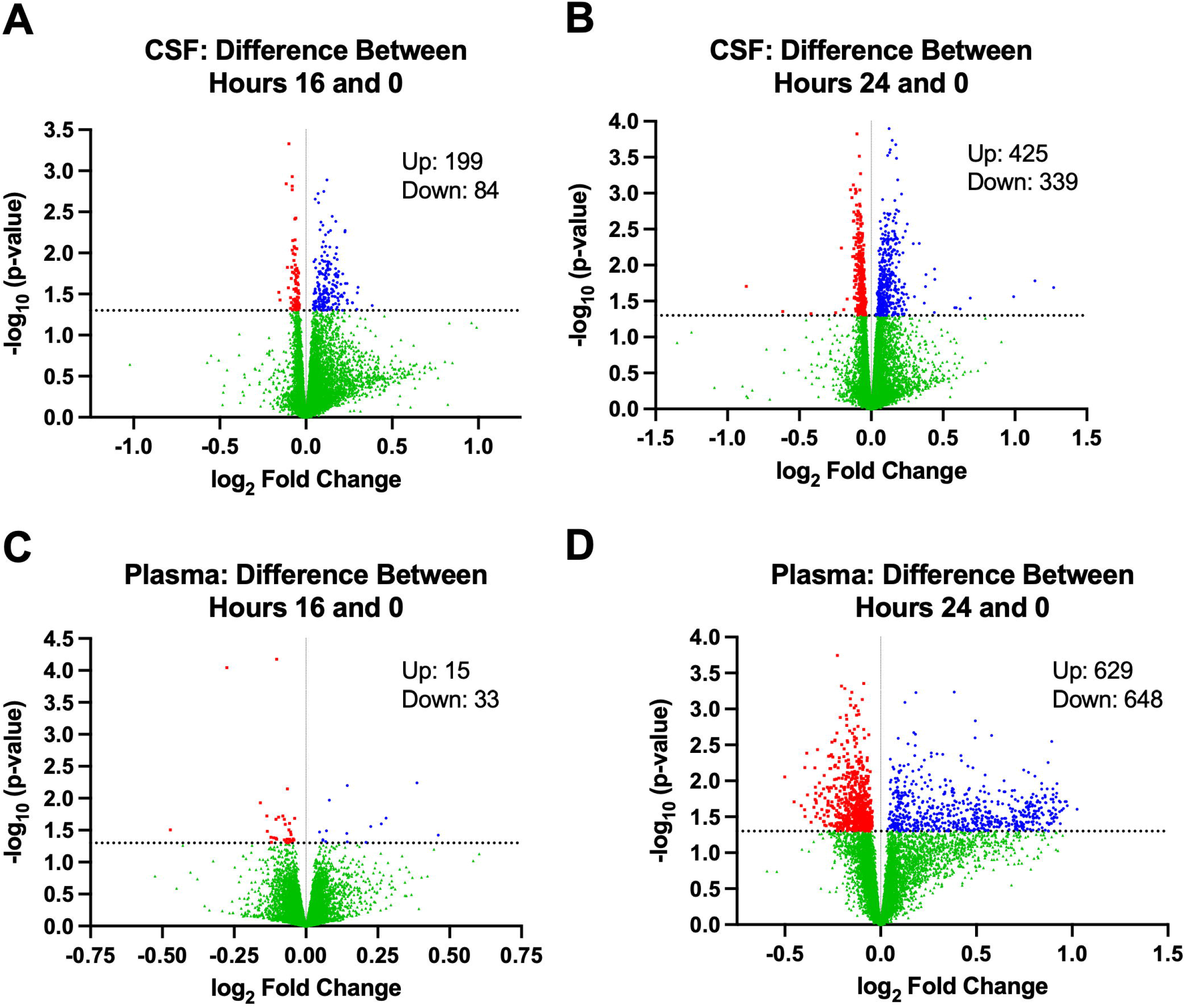
Suvorexant alters individual CSF and plasma protein levels. Volcano plots of SomaScan 11K differential expression for suvorexant 20 mg versus placebo, each expressed as change from hour 0: (A) CSF, hour 16 versus hour 0; (B) CSF, hour 24 versus hour 0; (C) plasma, hour 16 versus hour 0; (D) plasma, hour 24 versus hour 0. The x-axis shows log₂ fold change in the suvorexant group relative to placebo and the y-axis −log₁₀ of the unadjusted p-value from two-sided t-tests; the horizontal dashed line marks p = 0.05. Red, decreased p < 0.05; blue, increased p < 0.05; green, p ≥ 0.05. The number of increased and decreased proteins is inset in each panel. No protein survived Benjamini–Hochberg FDR correction in any contrast.

Given that no individual proteins were significant after FDR-adjustment, we tested group differences in pathways at hour 16 and 24 compared to baseline in two complementary analyses. To visualize the data and provide additional biological context for the changes associated with suvorexant compared to placebo at each time point, pathway and process enrichment analysis was performed through Metascape for up- and down-regulated pathways, protein-protein interactions, and Molecular Complex Detection (MCODE) subnetworks. During this exploratory analysis, pathways were included if their unadjusted p-value was < 0.05. Gene set enrichment analysis by the GAGE method^13^ compared the mean log fold change (logFC) of expressed genes in each pathway with the background genes across three databases: Gene Ontology (GO)-Biological Processes, GO-Molecular Function, and Kyoto Encyclopedia of Genes and Genomes (KEGG) Signaling and Metabolism. Then, the resulting enrichment statistic and its p-value were adjusted for multiple comparisons within each database, time point, and compartment. All pathways in CSF and plasma that were significant after FDR correction are shown in Supplementary Tables 1 and 2. Since GO terms have gene sets that overlaps, the number of significant terms exceeds the number of independent signals.

### Suvorexant Alters Different Protein Pathways in CSF and Plasma

#### CSF – Hour 16

Exploratory Metascape analysis of proteins with increased abundance in CSF in the suvorexant group at hour 16 compared to placebo identified clusters corresponding to cytoskeletal remodeling and cell motility (actin filament-based processes, regulation of lamellipodium organization, and spindle localization), synaptic and neuronal plasticity (modification of postsynaptic structure and synaptic vesicle endocytosis), immune cell trafficking and neuroinflammatory regulation (regulation of leukocyte cell-cell adhesion, positive regulation of leukocyte migration), and receptor-mediated signaling cascades (enzyme-linked receptor protein signaling pathway, regulation of G protein-coupled receptor signaling pathway) (Figure 4; Supplementary Figure 4). The protein-protein interaction network showed functional groupings with densely connected hubs corresponding to the cytoskeletal, synaptic, and receptor-signaling clusters. Four MCODE subnetworks were also identified: a GPCR-signaling module, an actin-regulatory module, a translation and RNA-processing module, and a cell-cycle-associated module. Among decreased enrichment pathways in CSF at hour 16 (Figure 5), identified clusters included humoral immune response, enzyme-linked receptor protein signaling pathway, regulation of angiogenesis, and miRNA targets of extracellular matrix and membrane receptors. There were too few significant pathways to perform MCODE analysis of proteins with decreased enrichment.

**Figure 4.**
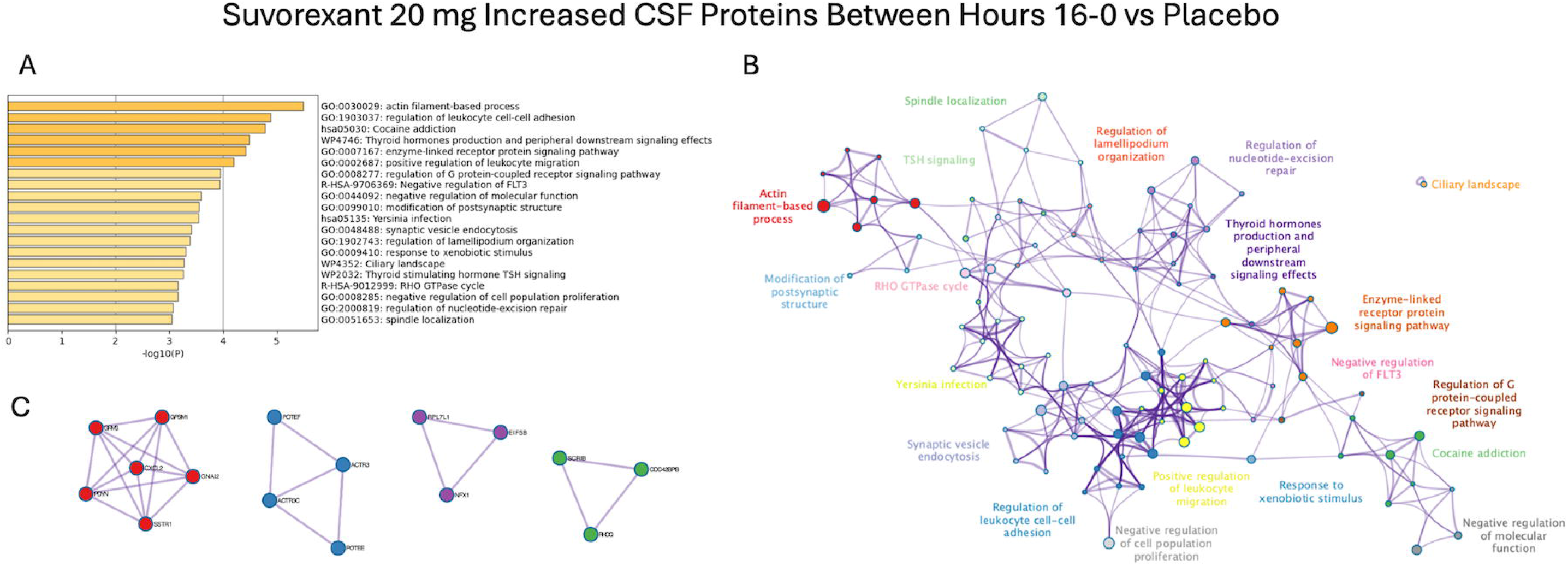
Metascape enrichment analysis of CSF proteins increased in the suvorexant 20 mg group compared to the placebo group at hour 16 relative to hour 0. (A) Top enriched ontology terms ranked by −log₁₀(P). (B) Network of enriched terms in which each node is a term, node size is proportional to the number of input proteins annotated to that term, and node color indicates cluster membership with cluster labels printed in the matching color. (C) MCODE subnetworks identified from the STRING and BioGRID protein–protein interaction network built from the same input list; each densely connected component is shown in a distinct color with nodes labeled by gene symbol. Four modules were identified, corresponding to G protein-coupled receptor signaling, actin regulation, translation and RNA processing, and cell cycle. Terms clustered into cytoskeletal remodeling and cell motility, synaptic and neuronal plasticity, immune cell trafficking, and receptor-mediated signaling cascades. Input proteins were those with unadjusted p < 0.05 in the SomaScan limma contrast.

**Figure 5.**
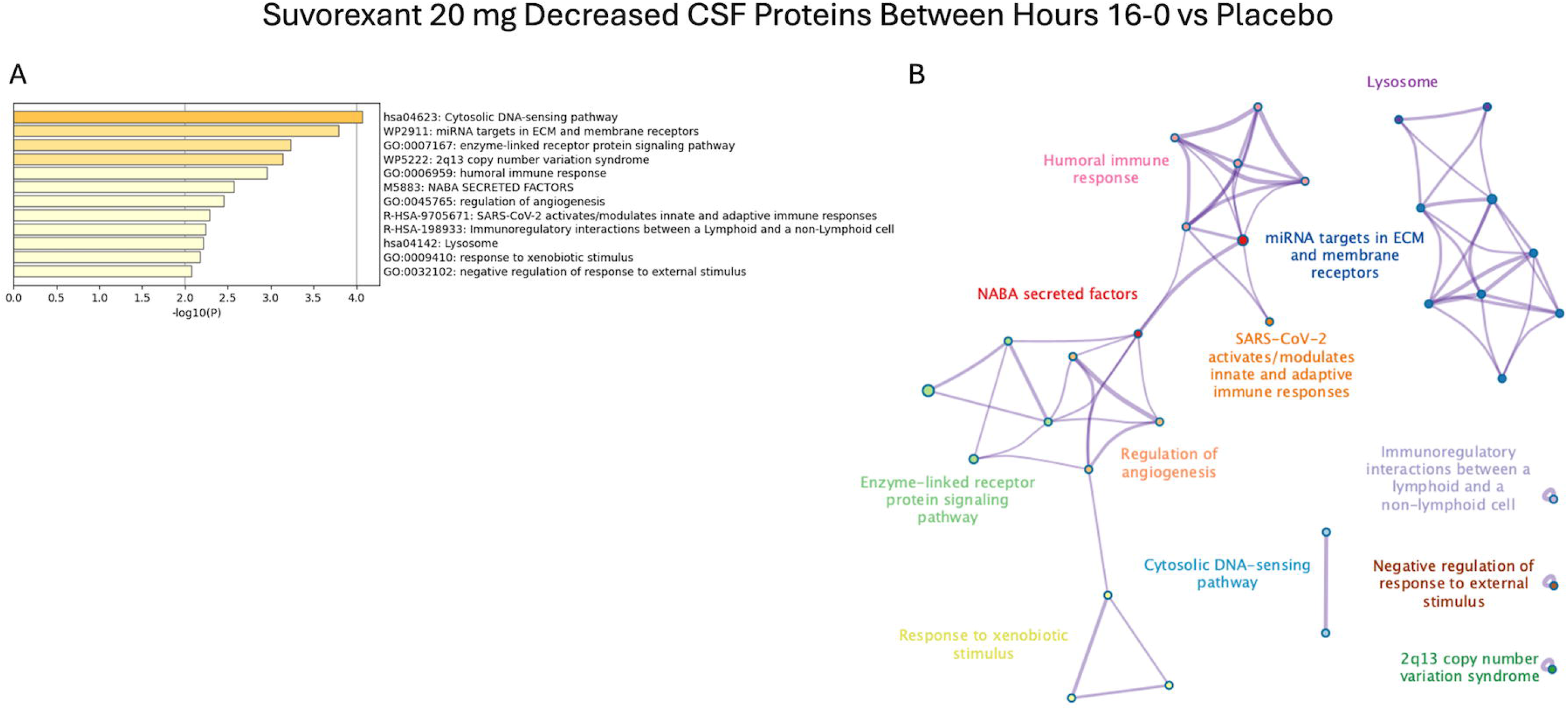
Metascape enrichment analysis of CSF proteins decreased in the suvorexant 20 mg group compared to the placebo group at hour 16 relative to hour 0. (A) Top enriched ontology terms ranked by −log₁₀(P). (B) Enrichment network with terms clustered into innate immune sensing and inflammatory signaling, extracellular matrix remodeling and vascular regulation, and receptor signaling and cellular homeostasis. Too few terms reached significance to support MCODE subnetwork analysis.

After correction for multiple comparisons, 8 GO-Biological Process terms, 24 GO-Molecular Function terms, and 1 KEGG pathway were FDR < 0.05 (Supplementary Table 1). All were enriched among proteins increased in the suvorexant group. The most significant terms were RNA related such as RNA binding (GO:0003723), mRNA binding (GO:0003729), translation (GO:0006412), and others (see Supplementary Table 1). These enrichment terms are not independent and have marked overlap. Multiple terms related to structural integrity and cytoskeletal dynamics were increased: cytoskeleton organization (GO:0007010), cellular component biogenesis (GO:0044085), organelle organization (GO:0006996), cellular component assembly (GO:0022607), and actin filament-based process (GO:0030029). These enrichment terms overlap and indicate that suvorexant is associated with greater abundance of proteins involved in the maintenance and remodeling of cellular architecture in CSF. Cadherin binding (GO:0045296), cytoskeletal protein binding (GO:0008092), phosphatidylinositol binding (GO:0035091), and ligase activity (GO:0016874) were also significantly enriched.

The endocytosis pathway (hsa04144) was the sole KEGG pathway to reach significance. There are several potential explanations for enrichment of the endocytosis pathway. First, neurons and glial cells rely on endocytosis for synaptic vesicle recycling, neurotransmitter receptor turnover, and clearance of cell-surface proteins.^14, 15, 16^ Enrichment suggests active membrane remodeling within the CNS. Second, exosomes originate from multivesicular endosomes,^17^ therefore enrichment of endocytic machinery may be associated with increased production of extracellular vesicles released into CSF^18^ by choroid plexus epithelium^19^ and neurons.^20^ Third, altered endosomal-lysosomal pathways are observed in neurodegenerative diseases where clearance is impaired including Alzheimer disease, Parkinson disease, and amyotrophic lateral sclerosis.^21, 22, 23^ Fourth, enrichment of endocytosis may also reflect altered choroid plexus or blood-CSF barrier transport activity. Although plausible, each of these mechanisms needs further study in preclinical models and human participants to confirm.

#### CSF – Hour 24

Metascape enrichment analysis of CSF proteins increased in the suvorexant group at hour 24 compared to placebo found clusters involving immune trafficking and inflammatory signaling (positive regulation of leukocyte migration, regulation of humoral immune response), vascular and endothelial processes (angiogenesis, VEGFA VEGFR2 signaling), and intracellular signaling and proteostasis pathways (regulation of catalytic activity, regulation of proteolysis) (Figure 6; Supplementary Figure 5). MCODE subnetworks included cell-polarity and adhesion, cell cycle, integrin-cytoskeletal, chemokine-cytokine, nucleolar/metabolic, and Rho-GTPase signaling modules. After FDR correction, however, a single GO-molecular function term remained significant at this time point: RNA binding (GO:0003723).

**Figure 6.**
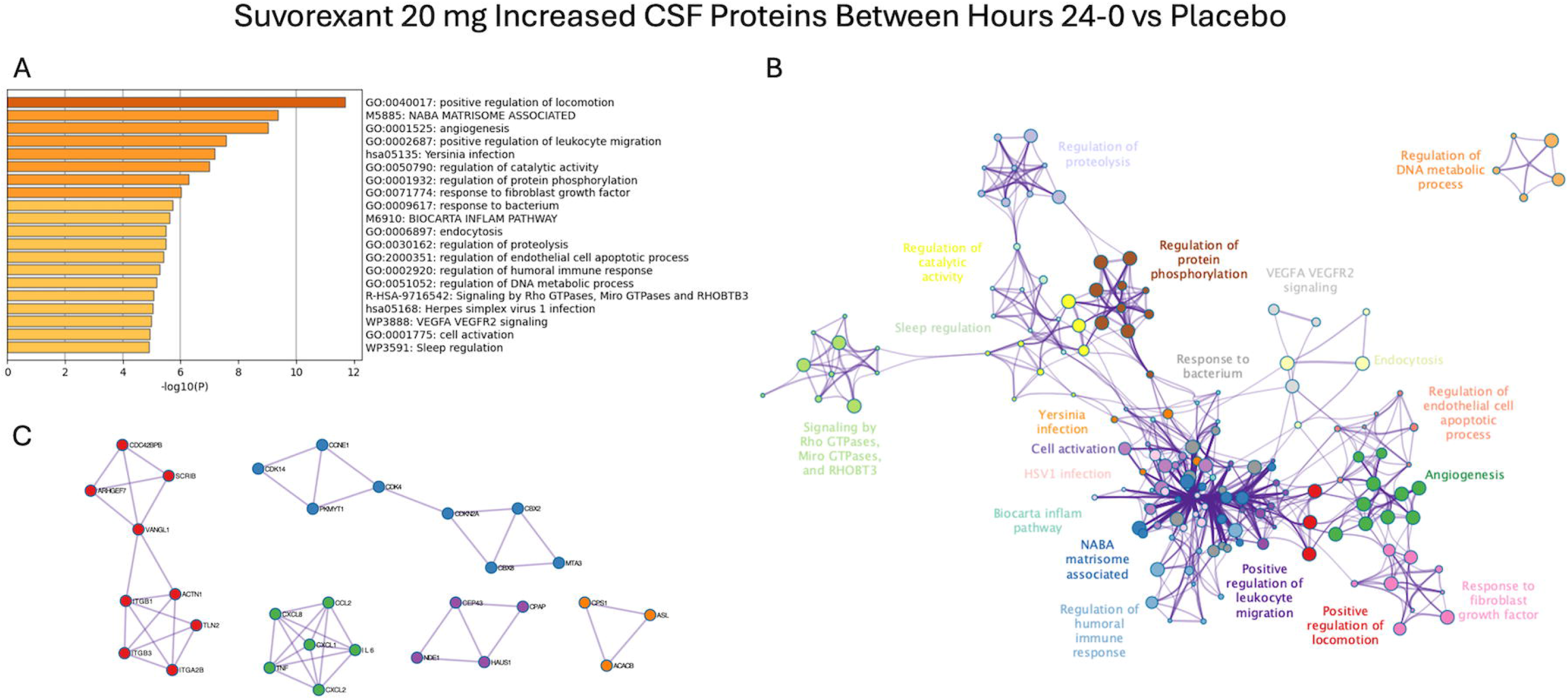
Metascape enrichment analysis of CSF proteins increased in the suvorexant 20 mg group compared to the placebo group at hour 24 relative to hour 0. (A) Top enriched ontology terms ranked by −log₁₀(P). (B) Enrichment network. (C) MCODE subnetworks corresponding to cell polarity and adhesion, cell cycle, integrin– cytoskeletal, chemokine–cytokine, nucleolar and metabolic, and Rho GTPase signaling modules.

Of the CSF proteins decreased at hour 24, Metascape identified clusters associated with receptor-driven proliferative and cytotoxic responses (enzyme-linked receptor protein signaling pathway, cell killing, and cell activation), cytokine-mediated immune cell effector functions and phagocytic clearance (phagocytosis, cellular response to cytokine stimulus), and tissue morphogenesis and vascular remodeling (Figure 7; Supplementary Figure 6). MCODE subnetworks included collagen-integrin adhesion, MHC/antigen-presentation, interleukin-receptor signaling, chromatin-remodeling, chemokine module, and cell cycle modules. No pathway was significantly decreased in CSF after adjustment for multiple comparisons at hour 24. Therefore, suvorexant increased 34 FDR-significant terms in CSF overall with only one significant term at hour 24.

**Figure 7.**
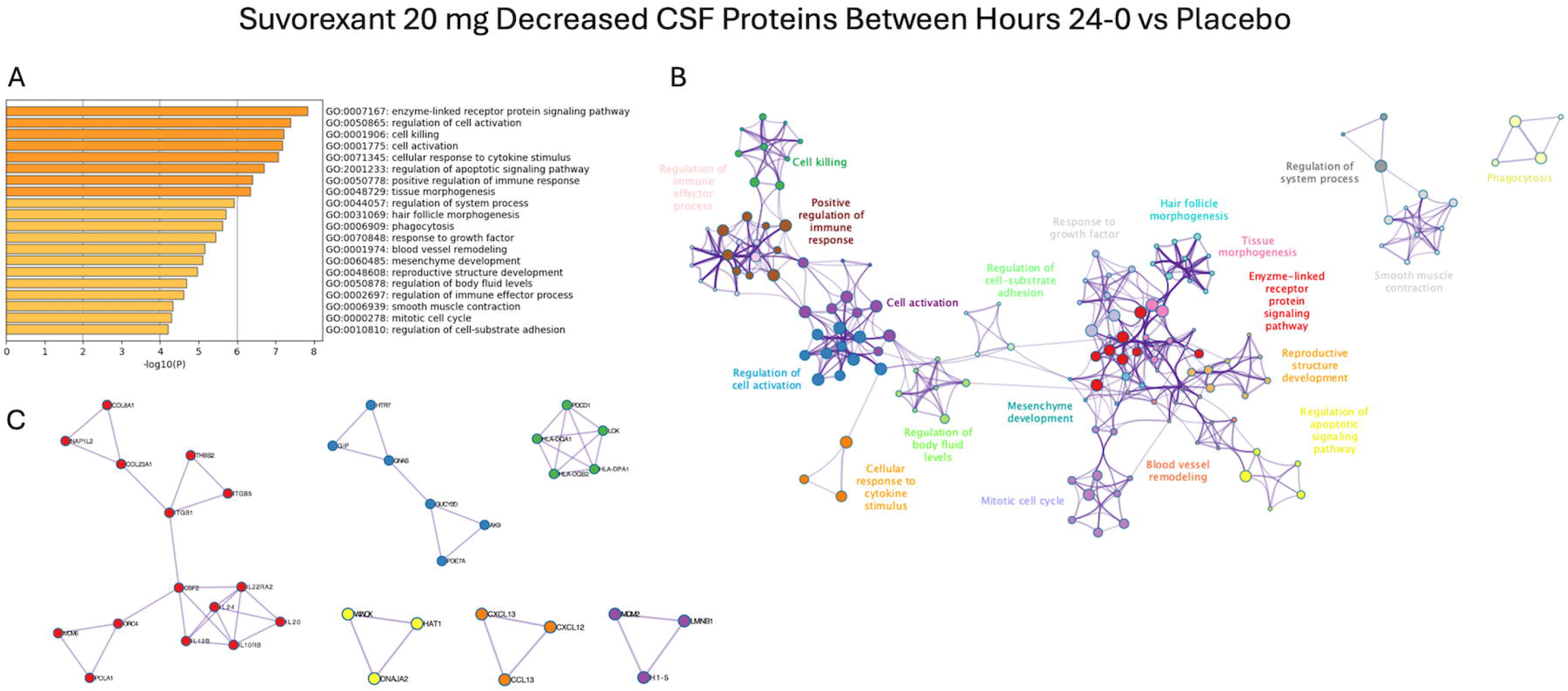
Metascape enrichment analysis of CSF proteins decreased in the suvorexant 20 mg group compared to the placebo group at hour 24 relative to hour 0. (A) Top enriched ontology terms ranked by −log₁₀(P). (B) Enrichment network. (C) MCODE subnetworks corresponding to collagen–integrin adhesion, MHC and antigen presentation, interleukin receptor signaling, chromatin remodeling, chemokine, and cell cycle modules.

#### Plasma – Hour 16

There were insufficient enriched pathways to perform Metascape analysis in plasma at hour 16. However, 12 GO-Biological Process terms, 1 GO-Molecular Function term, and 9 KEGG pathways were FDR < 0.05 at this time point (Supplementary Table 2). The most significantly enriched terms in the suvorexant group in plasma at hour 16 compared to placebo were organic acid metabolic process (GO:0006082), oxoacid metabolic process (GO:0043436), and carboxylic acid metabolic process (GO:0019752). The constituent proteins in these pathways are highly overlapping and represent a single signal. Organic acid catabolic process (GO:0016054) and carboxylic acid catabolic process (GO:0046395), which have identical gene sets, were also significantly enriched. These findings may indicate: 1) upregulation of proteins involved in the synthesis, transformation, and catabolism of organic and carboxylic acids in the plasma of suvorexant-treated participants; 2) release of these proteins in plasma that are not used or needed.

Small molecule catabolic process (GO:0044282) and small molecule metabolic process (GO:0044281) associated with the turnover of low-molecular-weight metabolites were both enriched in the suvorexant group compared to placebo. Multiple amino acid metabolic pathways reached significance as well, such as alpha-amino acid metabolic process (GO:1901605), L-amino acid metabolic process (GO:0170033), proteinogenic amino acid metabolic process (GO:0170039), and amino acid metabolic process (GO:0006520). Increased valine, leucine, and isoleucine degradation (hsa00280) and glycine, serine, and threonine metabolism (hsa00260) further support broad upregulation of amino acid catabolic and metabolic flux following suvorexant treatment. Carbon metabolism (hsa01200), pyruvate metabolism (hsa00620, fatty acid degradation (hsa00071), and metabolic pathways (hsa01100) were also significant. Peroxisome (hsa04146), an organelle central to fatty acid β-oxidation and reactive oxygen species metabolism, was enriched as was oxidoreductase activity (GO:0016491), which encompasses proteins that catalyze oxidation–reduction reactions.

In contrast with the CSF findings, the endocytosis pathway (hsa04144) exhibited significantly lower abundance in the suvorexant group relative to placebo at hour 16. While metabolic and catabolic pathways were broadly enriched, cellular vesicular uptake processes were relatively suppressed in plasma at 16 hours following suvorexant administration.

#### Plasma – Hour 24

In plasma at hour 24, Metascape enrichment analysis found an increased cluster in the suvorexant group compared to placebo involving cellular metabolic reprogramming with nucleoside phosphate metabolic process and carboxylic acid metabolic process as the top two terms (Figure 8; Supplementary Figure 7). A second cluster focused on protein homeostasis and intracellular degradation (regulation of translation, endocytosis, and autophagy). Others implicated vascular, immune, and stress-response signaling (VEGFA-VEGFR2 signaling, membrane trafficking, response to oxidative stress). MCODE subnetwork modules included a large ribosomal/translational module, nucleotide-metabolism, carboxylic acid metabolism, and cellular stress modules, and smaller nodes for VEGFA-VEGFR2 signaling, neutrophil degranulation, and oxidative stress response.

**Figure 8.**
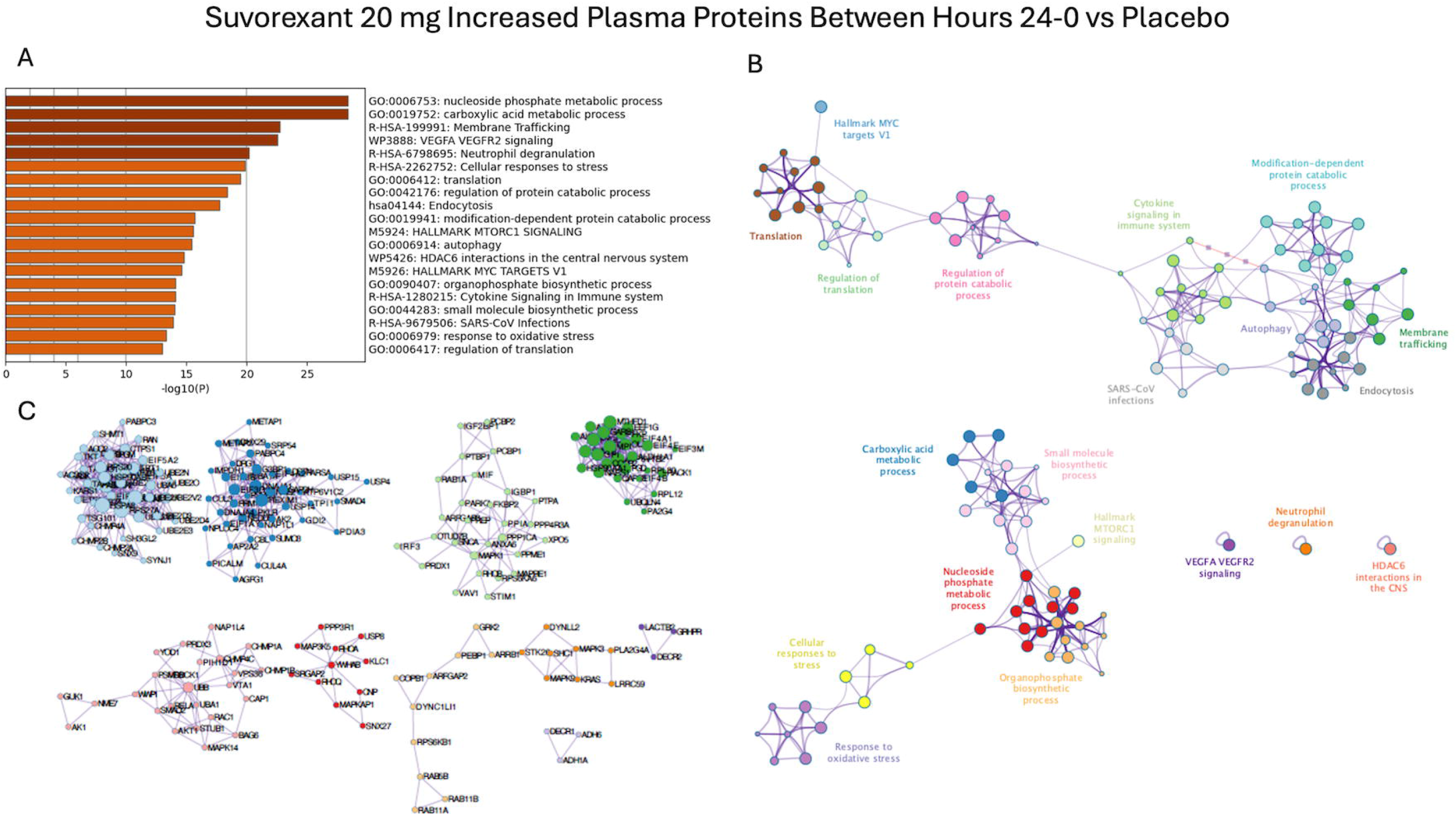
Metascape enrichment analysis of plasma proteins increased in the suvorexant 20 mg group compared to the placebo group at hour 24 relative to hour 0. (A) Top enriched ontology terms ranked by −log₁₀(P). (B) Enrichment network. (C) MCODE subnetworks including a large ribosomal and translational module together with nucleotide metabolism, carboxylic acid metabolism, and cellular stress modules and smaller VEGFA–VEGFR2 signaling, neutrophil degranulation, and oxidative stress response modules.

Of the proteins decreased in plasma at hour 24 in the suvorexant group compared to placebo, Metascape identified matrisome secretion, epithelial-to-mesenchymal transition, and receptor-tyrosine kinase cascades, a developmental and morphogenetic cluster including vasculature development and tissue morphogenesis, and a cluster implicated in immune regulation and inflammatory signaling (Figure 9; Supplementary Figure 8). The protein– protein interaction network organized the terms into a large central hub connecting cell activation, immune system development, cytokine signaling, and multicellular organismal-level homeostasis, with branches to enzyme-linked receptor protein signaling, vasculature development, NABA core matrisome, neuron projection development, cell–cell adhesion, and Hallmark epithelial mesenchymal transition. MCODE subnetworks included collagen-extracellular-matrix, Wnt developmental signaling, cell-surface receptor, MHC/immune-recognition, and G-protein signaling modules.

**Figure 9.**
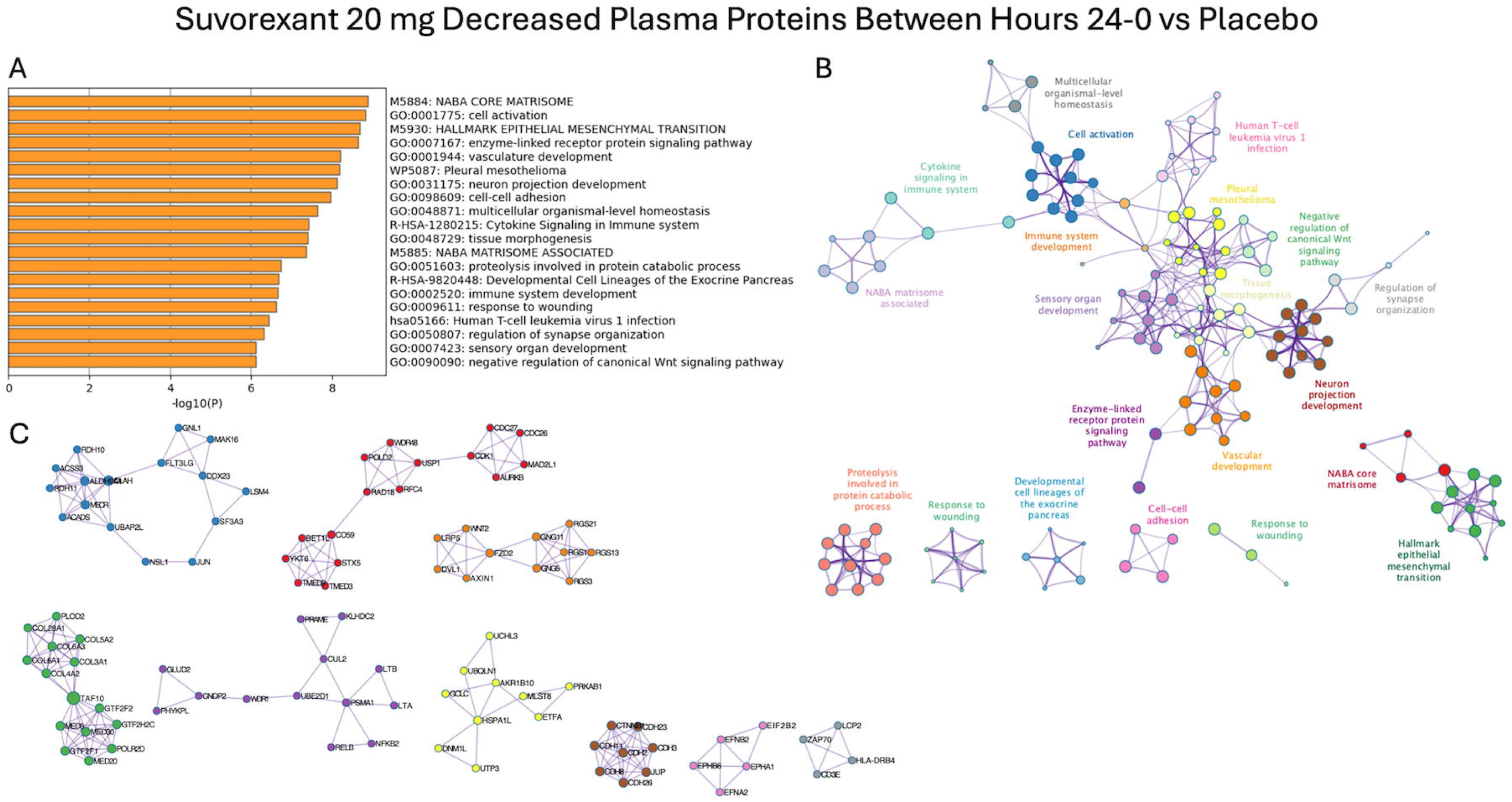
Metascape enrichment analysis of plasma proteins decreased in the suvorexant 20 mg group compared to the placebo group at hour 24 relative to hour 0. (A) Top enriched ontology terms ranked by −log₁₀(P). (B) Enrichment network. The network organized into a large central hub connecting cell activation, immune system development, cytokine signaling, and multicellular organismal-level homeostasis, with branches to enzyme-linked receptor protein signaling, vasculature development, NABA core matrisome, neuron projection development, cell–cell adhesion, and Hallmark epithelial–mesenchymal transition. (C) MCODE subnetworks including collagen– extracellular matrix, Wnt developmental signaling, cell-surface receptor, MHC and immune recognition, and G protein signaling modules.

One hundred and thirty-five GO-Biological Process terms, 55 GO-Molecular Function terms, and 27 KEGG pathways were FDR < 0.05 in plasma at hour 24 in the suvorexant group compared to placebo. Multiple pathways linked to translation were significantly enriched in participants receiving suvorexant including: translation (GO:0006412), translational initiation (GO:0006413), regulation of translational initiation (GO:0006446), and others. Processes related to upregulation of ribonucleoprotein complexes, such as RNA binding (GO:0003723) and mRNA binding (GO:0003729), were also significant. Multiple interconnected pathways related to nucleotide biosynthesis and catabolism (nucleobase-containing small molecule metabolic process (GO:0055086), nucleotide metabolic process (GO:0009117), nucleotide biosynthetic process (GO:0009165), ATP binding (GO:0005524), nucleotide metabolism (hsa01232)) were also enriched. Additional pathways reflecting intracellular protein degradation, such as catabolic process (GO:0009056), proteolysis involved in protein catabolic process (GO:0051603), protein catabolic process (GO:0030163), and others, were increased. The ubiquitin-proteasome system was implicated through ubiquitin-dependent protein catabolic process (GO:0006511), ubiquitin-like protein ligase binding (GO:0044389), and ubiquitin protein ligase binding (GO:0031625). Autophagic pathways were also increased with enrichment of macroautophagy (GO:0016236), autophagy (GO:0006914), autophagosome maturation (GO:0097352), and autophagy – animal (hsa04140).

A cluster of vesicular transport pathways showed significantly increased abundance. Endosome organization (GO:0007032), vesicle-mediated transport (GO:0016192), multivesicular body assembly (GO:0036258), endocytosis (hsa04144), among others, were enriched. Interestingly, endocytosis was decreased in plasma while increased in CSF at hour 16 but increased in plasma at hour 24. There were also multiple cytoskeleton remodeling pathways upregulated. Cytoskeleton organization (GO:0007010), actin filament organization (GO:0007015), microtubule cytoskeleton organization (GO:0000226), and several others were significant. Finally, protein quality control pathways were significantly enriched, including protein folding (GO:0006457), chaperone-mediated protein folding (GO:0061077), and response to heat (GO:0009408).

Several intracellular signaling cascades were positively enriched: mTOR signaling pathway (hsa04150), insulin signaling pathway (hsa04910), neurotrophin signaling pathway (hsa04722), ErbB signaling pathway (hsa04012), VEGF signaling pathway (hsa04370), sphingolipid signaling pathway (hsa04071), B cell receptor signaling pathway (hsa04662), and G protein activity (GO:0003925). These gene sets overlap substantially. Mitophagy (hsa04137), protein processing in endoplasmic reticulum (hsa04141), glutathione metabolism (hsa00480), purine metabolism (hsa00230), nucleotide metabolism (hsa01232), and several metabolic pathways (metabolic pathways (hsa01100), carbon metabolism (hsa01200), pyruvate metabolism (hsa00620), biosynthesis of amino acids (hsa01230)) were also significant. Tau protein binding (GO:0048156) and phosphoprotein binding (GO:0051219) reached significance among molecular function terms.

Thirteen terms were significantly decreased in the suvorexant group in plasma at hour 24 and comprise two groups. The first is a receptor and cytokine signaling group: cytokine - cytokine receptor interaction (hsa04060) was the most strongly suppressed KEGG pathway, followed by neuroactive ligand-receptor interaction (hsa04080), signaling receptor activity (GO:0038023), molecular transducer activity (GO:0060089), transmembrane signaling receptor activity (GO:0004888), signaling receptor regulator activity (GO:0030545), receptor ligand activity (GO:0048018), signaling receptor activator activity (GO:0030546), and cytokine activity (GO:0005125). The second is a proteolytic and matrix group: serine-type peptidase activity (GO:0008236) and serine hydrolase activity (GO:0017171) were both significantly decreased, as were protein digestion and absorption (hsa04974) and extracellular matrix structural constituent (GO:0005201). The direction of the immune signal at this time point is therefore mixed rather than uniformly increased: circulating cytokine and receptor-ligand signaling was suppressed, while the intracellular Fc-receptor and antigen-receptor effector modules noted above were modestly enriched and sit among the weakest significant KEGG terms.

### Cross-Compartment Pathway Enrichment

Thirty-two GO and KEGG pathways achieved FDR < 0.05 in both CSF and plasma. All of them were significant in CSF at hour 16 and in plasma at hour 24 with only 1 pathway reaching significance in both compartments at the same timepoint (RNA Binding). All 24 CSF molecular function terms recur in plasma at hour 24, as do 7 of the 8 biological process terms; only mRNA metabolic process (GO:0016071) fails to recur. Endocytosis (hsa04144) was the sole KEGG term in this shared set. RNA binding (GO:0003723) was the only term significant in three of the four comparisons (CSF hour 16, CSF hour 24, plasma hour 24).

## DISCUSSION

This study found that suvorexant increases plasma pT181/T181 and peaks approximately 15 hours post-dose. In contrast, we previously reported that suvorexant decreases CSF pT181/T181. In CSF, every FDR-significant pathway at both hours 16 and 24 was enriched among proteins that increased in response to orexin antagonism; no pathway was significantly decreased centrally at either time point. The CSF signature at hour 16 consisted of increased RNA and nucleotide binding, translation, cytoskeletal organization, cadherin and phosphatidylinositol binding, guanyl-nucleotide and G-protein modules, and KEGG endocytosis; at hour 24 only RNA binding remained significant. In plasma, hour 16 was dominated by hepatic and catabolic metabolism pathways, and hour 24 carried the great majority of the study’s significant terms, comprising translation and RNA handling, nucleotide metabolism, ubiquitin-proteasome and autophagic catabolism, endosomal and multivesicular body sorting, cytoskeletal organization and chaperone-mediated folding, together with a smaller set of significantly decreased terms covering cytokine and receptor-ligand signaling, serine protease activity and extracellular matrix constituents.

The divergent CSF and plasma pT181/T181 trajectories are consistent with altered intercompartmental handling of phosphorylated tau, but the present data cannot distinguish selective CNS-to-blood export from altered peripheral clearance, differential tau processing, assay-related effects, or compartment-specific changes in phosphorylation. We hypothesize that selective export from the central to the peripheral compartment is one mechanism to explain the increase in plasma pT181/T181. The absence of a significant effect on plasma Aβ40, Aβ42, or unphosphorylated T181 and T217 argues against a simple increase in bulk transfer of CNS-derived species into blood since a clearance effect of that kind should also affect Aβ and unphosphorylated tau. Further, the CSF pool of tau is approximately 20-fold more concentrated than the plasma pool.^24^ Therefore, exporting a smaller relative fraction of CSF p-tau would be predicted to increase the plasma p-tau pool by a greater relative amount. Proteins are cleared from CSF by several routes: bulk outflow via glymphatic exchange along perineural and dural lymphatic pathways to cervical lymph nodes and blood,^25, 26, 27, 28^ receptor-mediated transport at the blood-CSF and blood-brain barriers for specific proteins,^29, 30^ and local proteolytic degradation.^31^ However, our data cannot measure any of these processes directly.

There are alternative hypotheses, however, including reduced peripheral proteolytic capacity for phosphorylated tau and/or central versus peripheral differences in kinase engagement with tau. Both alternatives are difficult to assess since the proteomic data does not measure activity or concentration, although pathways that would support these hypotheses also do not align with the biomarker changes. For example, tau is a substrate for circulating serine proteases,^32^ and phosphorylation prevents cleavage with paired helical filament tau unless first dephosphorylated.^33^ A fall in circulating serine protease capacity would therefore lengthen the residence time of the phosphorylated species preferentially, raising the measured ratio of phosphorylated to unphosphorylated T181 without any kinase acting on tau in blood. Although serine-type peptidase activity (GO:0008236) and serine hydrolase activity (GO:0017171) were both significantly decreased in plasma in the suvorexant group, this occurred at hour 24 rather than hour 16, when group differences in pT181/T181 were greatest. Multiple pathways associated with the ubiquitin-proteasome-chaperone system that degrades tau were also enriched at hour 24 in plasma but not at hour 16 or in CSF: modification-dependent protein catabolic process (GO:0019941), ubiquitin-dependent protein catabolic process (GO:0006511), regulation of proteasomal protein catabolic process (GO:0061136), and others.

DORAs, such as suvorexant and lemborexant, reduce pT181/T181 in human CSF^12^ and tau phosphorylation in transgenic mice.^10^ In mice, lemborexant decreased phosphorylated tau and tau-mediated neurodegeneration via protein kinase A (PKA) and microglia changes.^10^ The CSF signature, however, does not contain pathways directly associated with tau phosphorylation that reached FDR < 0.05 at either time point. In contrast to CSF, multiple pathways potentially related to tau phosphorylation were enriched in plasma at hour 24 but not hour 16. KEGG pathways, for example, containing the tau kinases were increased:^34, 35^ neurotrophin signaling (hsa04722), ErbB (hsa04012), vascular endothelial growth factors (VEGF) (hsa04370), mammalian target of rapamycin (mTOR) (hsa04150)^36^ and insulin signaling (hsa04910). The neurotrophin signaling pathway includes extracellular signal-regulated kinase (ERK) kinases that phosphorylate tau at threonine-181.^37, 38^ Again, the timing of these proteomic changes does not align with the group differences in pT181/T181.

This study represents one of the first attempts to measure the effect of DORAs on plasma AD biomarkers and to map the proteomic effects of orexin antagonism across CSF and plasma in cognitively unimpaired adults. However, there are several important limitations. Suvorexant-mediated changes were measured over hours in a small cohort of cognitively unimpaired middle-aged adults. We identified several pathway-level changes significant after FDR-correction despite the limited power to detect individual protein changes. SomaScan and NULISA measure relative protein abundance, not activity or concentration, and both assays assess the extracellular compartment, so terms describing intracellular processes reflect the origin and annotation of the proteins detected rather than processes occurring in CSF or plasma. Further, measurement of additional forms of phosphorylated tau in CSF and plasma as well as confirmation of proteomic changes with dedicated mass spectrometry or immunoassays are needed. We emphasize that no published work connects orexin signaling to all of the mechanisms discussed and that the link from suvorexant to these plasma changes is inferred from the enrichment pattern and needs further testing. Given the rapid adoption of plasma AD biomarkers for clinical care and research, factors that influence their levels, such as medications like suvorexant, must be identified and their effects on interpretation understood.

## METHODS

### Participant Characteristics and Study Design

CSF and plasma samples collected from twenty-five participants aged 45 to 65 years in a previously published randomized placebo-controlled trial were included in the study.^12^ Inclusion criteria included a Mini-Mental State Examination (MMSE) score ≥27 (i.e., cognitively unimpaired),^39^ absence of neurological or sleep disorders, and actigraphic sleep efficiency < 85%. Participants were randomized to placebo (N=13) and suvorexant 20 mg (N=12). There was also a suvorexant 10 mg group which was not included in our analyses. Participant characteristics are shown in Table 1. The study was conducted at the Washington University School of Medicine in St Louis, Missouri. The study protocol was approved by the Washington University Institutional Review Board. The Clinical Trials number was NCT03077620. All participants completed written informed consent and were compensated for their participation in the study.

**Table 1:** Participant Characteristics.

|  | Placebo<br>(N=13) | Suvorexant 20 mg<br>(N=12) |
|---|---|---|
| Age (years), mean (SD) | 55.94 (6.10) | 54.30 (5.66) |
| Sex (%, N) | F: 61.5%, 8<br>M: 38.5%, 5 | F: 75%, 9<br>M: 25%, 3 |
| Race (%), N | AA: 23.1%, 3<br>C: 76.9%, 10 | AA: 8.3%, 1<br>C: 91.7%, 11 |
| ApoE4+ (%), N | 23.1%, 3 | 50%, 6 |
| MMSE, mean (SD) | 29.38 (0.87) | 29.67 (0.49) |
| Screening actigraphic sleep efficiency (%), mean (SD) | 72.96% (9.36) | 78.82 (6.29) |
| Screening actigraphic total sleep time (min), mean (SD) | 361.49 (59.58) | 379.36 (38.55) |

SD: standard deviation; ApoE4: apolipoprotein E4; MMSE: Mini-Mental Status Examination; min: minutes; F: female; M: male; AA: African-American; C: Caucasian

Participants were admitted to the Clinical Translational Research Unit (CTRU). Both an intravenous catheter (IV) and an intrathecal lumbar catheter were placed. Collection of paired CSF and blood samples started in all participants at 20:00. Participants, research staff, and laboratory personnel were blinded to treatment status. CSF was collected every 2 hours for 36 hours and blood collected at the following hours was used in the analyses: 0, 6, 8, 12, 16, 20, 24, 28, 32, and 36. CSF and blood samples were processed as previously described and stored at -80°C until analysis.^12, 40^ After acclimation to the lumbar catheter and collection of the initial samples, participants received their first dose of placebo or suvorexant 20 mg at 21:00 (hour 1). The participants received their second dose of the same intervention from the first night at 21:00 (hour 25) on the second night. The IV and lumbar catheter were removed on day 3 at 08:00. Participants had meals served at 09:00, 13:00, and 18:00. Polysomnography was performed as previously reported throughout each participant’s admission to the CTRU.

### Alzheimer Disease Biomarker Quantification

All CSF samples were processed and measured for CSF Aβ40, Aβ42, T181, pT181, T217, and phosphorylated T217 (pT217) as previously described.^12^ Plasma Aβ40, Aβ42, T181, pT181, T217, and pT217 concentrations were quantified by C2N Diagnostics, LLC (St. Louis, MO) using the immunoprecipitation liquid chromatography-tandem mass spectrometry (LC-MS/MS) assays.^41, 42, 43^ Plasma pT217 was below the limit of quantitation in 64.6% of samples and was not included in the analysis. Plasma tau and p-tau could not be quantified in one placebo participant at hour 0 and this participant was excluded from the analyses.

### SomaScan Assay

Human CSF and plasma samples from participants treated with suvorexant 20 mg or placebo from t=0 (20:00 on Day 1), t=16 (12:00 on Day 2), and t=24 (20:00 on Day 2) timepoints were assayed using the SomaScan 11K kit (SomaLogic, Boulder, CO), which is a high-scale aptamer-based proteomic platform that measures relative concentrations of ∼11,000 proteins. These timepoints were chosen to capture both the greatest AD biomarker change (hour 16) and circadian-matched comparisons (hour 24) relative to baseline. CSF and plasma samples were available from 13 participants in the placebo group and 12 participants in the suvorexant group at hours 0 and 16. At hour 24, there was one participant in each group without CSF available for analysis. All CSF and plasma samples were analyzed per manufacturer’s instructions. Briefly, we input 50 μL of plasma samples and prepared in 3 separate plate wells at 40%, 1%, and 0.005% dilution, respectively, and a single 20% dilution across all CSF sample wells on the SomaScan assay plate. Samples were hybridized into Agilent Surescan MicroArray slides and then scanned at a 5-μm resolution to detect CY3 fluorescence. Gridding and image analysis were done using Agilent Feature Extraction v10.7.3.1. Raw signals were standardized according to SomaScan.^44^

### NULISA

Ultra-sensitive quantification of 120 proteins in CSF and plasma was performed using NULISAseq CNS Disease Panel 120 (Alamar Biosciences, Fremont, CA), a bead-based multiplex immunoassay that amplifies detection signals via nucleic acid ligation and next-generation sequencing readout.^45^ Plasma samples were available from all 25 participants. CSF samples from 12 participants who received placebo and 11 participants who received suvorexant 20 mg were available for analysis. NULISA minimizes assay background and increases sensitivity at the attomole level. Ten µl of samples was used at each time point for each participant and concentration is reported as NULISA Protein Quantification (NPQ) units. NPQ units are the result of normalizing the sequence quantification counts by intraplate and intensity variability and then are log2-transformed to approximate normality.^46^

### Differential Abundance Analysis

Vendor-standardized SomaScan signal data were subjected to differential expression analysis using the limma package in R. For each compartment (CSF and plasma), two comparisons were performed: suvorexant 20 mg versus placebo at hour 16 versus baseline (T16 vs. T0) and at hour 24 versus baseline (T24 vs. T0). A participant-paired moderated t-test with empirical Bayes shrinkage of variance was used to identify differentially expressed proteins. The limma package generates both nominal p-values and multiple testing-corrected adjusted p-values using the Benjamini-Hochberg false discovery rate (FDR) procedure. Given that pathway-level genes tend to be co-regulated, proteins with nominal p < 0.05 were included in downstream pathway enrichment analyses to reduce false negative rates at the individual protein level.

### Network and Pathway Analysis

Upregulated and downregulated protein gene lists based on the SomaScan data were processed through the web interface Metascape for the gene ontology, biological function, and pathway analyses. The Metascape utilizes the well-adopted hypergeometric test to identify statistically enriched ontology terms. Significant terms were defined as those with a minimum count of three genes and an enrichment factor more than 1.5 (i.e., the ratio between the observed counts and the counts expected by chance) at p < 0.05 or Benjamini-Hochberg FDR corrected p < 0.05. The Metascape also uses Kappa similarity algorithm to absorb most redundancies of biological terms into representative clusters. Clusters were named based on the most statistically significant term within the cluster. Analysis in Metascape provided the top upregulated and downregulated gene ontology clusters with their representative enriched terms for illustration and discussion.

To capture the relationships between terms, we selected the top 15 enriched terms from each of the 20 top cluster terms and rendered them as a network plot visualized with Cytoscape. The terms with the best p-values had no more than 15 terms per cluster and no more than 250 terms in total. Circle node color is paired with the enriched terms that represent cluster identity (text written in the same color in the figure) and node size is proportional to the number of input genes included in the selected terms. Terms with a Kappa similarity score > 0.3 are linked by a branch whose thickness represents the similarity score. Finally, we use STRING and BioGrid to reveal enriched protein-protein physical interactions. Protein interaction networks containing between 3 and 500 proteins were included and analyzed through the MCODE algorithm to identify densely connected network components. The three best-scoring terms by p-value were retained as a functional description of the corresponding components.

### Statistics

Serial CSF Aβ, tau, and phospho-tau data were analyzed using general linear mixed models to account for dependencies among longitudinal measurements. The mixed model used a compound symmetry covariance matrix and was fit using restricted maximum likelihood (REML) as implemented in GraphPad Prism 8.0. Intervention group, time of day, and intervention × time interaction were treated as fixed effects; random intercepts and slopes accommodated individual variation. Plasma AD biomarkers (Aβ40, Aβ42, pT181, T181, T217) were analyzed using the same mixed model framework.

Two-sided t-tests were used to test for group differences in individual proteins measured by SomaScan and NULISA. For SomaScan proteomic analyses, differential expression was performed using the limma package in R with a participant-paired moderated t-test and empirical Bayes shrinkage of variance. Multiple testing correction was applied using the Benjamini-Hochberg FDR procedure. Individual proteins with nominal p < 0.05 were used for downstream pathway enrichment analyses. For pathway analyses, statistical significance was defined as FDR < 0.05. For the primary outcome, pT181/T181, significance was set at p < 0.05.

## Data Availability

All data produced in the present study are available upon reasonable request to the authors.

## Data Availability

Participant consent does not allow for open access sharing of data, but deidentified data is available to qualified investigators upon request.

## ACKNOWLEDGEMENTS

We thank the research participants for their time. We also appreciate Nicky “Cassandra” Cox at C2N Diagnostics for project management support. We acknowledge the use of artificial intelligence-assisted technologies (Claude, Anthropic) to improve clarity, spelling, and grammar of this manuscript. All edits were reviewed by the authors, who take full responsibility for the content of this article.

## AUTHOR CONTRIBUTIONS

Conceptualized the study: B.P.L.

Collected and managed the data: T.J.P., J.Y., B.P.L.

Performed formal analyses: T.J.P., J.Y., A.L., B.P.L.

Wrote the original draft: T.J.P., B.P.L.

Revised and edited the manuscript: T.J.P., J.Y., A.L., L.I., O.M.B., B.P.L.

## REFERENCES

1. Masters CL, Bateman R, Blennow K, Rowe CC, Sperling RA, Cummings JL. Alzheimer’s disease. Nat Rev Dis Primers 1, 15056 (2015).

2. Barthélemy NR, et al. Highly accurate blood test for Alzheimer’s disease is similar or superior to clinical cerebrospinal fluid tests. Nat Med 30, 1085–1095 (2024).

3. Rissman RA, et al. Plasma Aβ42/Aβ40 and phospho-tau217 concentration ratios increase the accuracy of amyloid PET classification in preclinical Alzheimer’s disease. Alzheimers Dement 20, 1214–1224 (2024).

4. Salvadó G, et al. Plasma Phosphorylated Tau 217 to Identify Preclinical Alzheimer Disease. JAMA Neurol 82, 1122–1134 (2025).

5. Janelidze S, et al. Plasma Phosphorylated Tau 217 and Aβ42/40 to Predict Early Brain Aβ Accumulation in People Without Cognitive Impairment. JAMA Neurol 81, 947–957 (2024).

6. McConnell BV, Deng Y, Lucey BP. Sleep and Neurodegeneration: Examining Potential Physiological Mechanisms. Curr Sleep Med Rep 11, 10.1007 (2025).

7. Pedersen TJ, et al. Wake-promoting neuromodulators in Alzheimer’s disease: Implications for sleep and brain clearance. Alzheimers Dement 22, e71298 (2026).

8. Li J, Hu Z, de Lecea L. The hypocretins/orexins: integrators of multiple physiological functions. Br J Pharmacol 171, 332–350 (2014).

9. Kang J-E, et al. Amyloid-β dynamics are regulated by orexin and the sleep-wake cycle. Science 326, 1005–1007 (2009).

10. Parhizkar S, et al. Lemborexant ameliorates tau-mediated sleep loss and neurodegeneration in males in a mouse model of tauopathy. Nat Neurosci 28, 1460–1472 (2025).

11. Michelson D, et al. Safety and efficacy of suvorexant during 1-year treatment of insomnia with subsequent abrupt treatment discontinuation: a phase 3 randomised, double-blind, placebo-controlled trial. Lancet Neurol 13, 461–471 (2014).

12. Lucey BP, et al. Suvorexant acutely decreases tau phosphorylation and Aβ in the human CNS. Ann Neurol 94, 27–40 (2023).

13. Luo W, Friedman MS, Shedden K, Hankenson KD, Woolf PJ. GAGE: generally applicable gene set enrichment for pathway analysis. BMC Bioinformatics 10, 161 (2009).

14. Saheki Y, De Camilli P. Synaptic vesicle endocytosis. Cold Spring Harbor Perspectives in Biology 4, a005645 (2012).

15. Anggono V, Huganir RL. Regulation of AMPA receptor trafficking and synaptic plasticity. Current Opinion in Neurobiology 22, 461–469 (2012).

16. Sheldon AL, González MI, Krizman-Genda EN, Susarla BTS, Robinson MB. Ubiquitination-mediated internalization and degradation of the astroglial glutamate transporter, GLT-1. Neurochemistry International 53, 296–308 (2008).

17. van Niel G, D’Angelo G, Raposo G. Shedding light on the cell biology of extracellular vesicles. Nature Reviews Molecular Cell Biology 19, 213–228 (2018).

18. Chiasserini D, et al. Proteomic analysis of cerebrospinal fluid extracellular vesicles: a comprehensive dataset. Journal of Proteomics 106, 191–204 (2014).

19. Balusu S, et al. Identification of a novel mechanism of blood-brain communication during peripheral inflammation via choroid plexus-derived extracellular vesicles. EMBO Molecular Medicine 8, 1162–1183 (2016).

20. Lachenal G, et al. Release of exosomes from differentiated neurons and its regulation by synaptic glutamatergic activity. Mol Cell Neurosci 46, 409–418 (2011).

21. Nixon RA. The role of autophagy in neurodegenerative disease. Nature Medicine 19, 983–997 (2013).

22. Simoes S, et al. Tau and other proteins found in Alzheimer’s disease spinal fluid are linked to retromer-mediated endosomal traffic in mice and humans. Science Translational Medicine 12, eaba6334 (2020).

23. Cuervo AM, Stefanis L, Fredenburg R, Lansbury PT, Sulzer D. Impaired degradation of mutant alpha-synuclein by chaperone-mediated autophagy. Science 305, 1292–1295 (2004).

24. Gonzalez-Ortiz F, et al. Brain-derived tau: a novel blood-based biomarker for Alzheimer’s disease-type neurodegeneration. Brain 146, 1152–1165 (2023).

25. Ma Q, Ineichen BV, Detmar M, Proulx ST. Outflow of cerebrospinal fluid is predominantly through lymphatic vessels and is reduced in aged mice. Nat Commun 8, 1434 (2017).

26. Smyth LCD, et al. Identification of direct connections between the dura and the brain. Nature 627, 165–173 (2024).

27. Proulx ST. Cerebrospinal fluid outflow: a review of the historical and contemporary evidence for arachnoid villi, perineural routes, and dural lymphatics. Cell Mol Life Sci 78, 2429–2457 (2021).

28. Iliff JJ, et al. A paravascular pathway facilitates CSF flow through the brain parenchyma and the clearance of interstitial solutes, including Amyloid β. Sci Transl Med 4, 147ra111 (2012).

29. Fujiyoshi M, et al. Amyloid-beta peptide(1-40) elimination from cerebrospinal fluid involves low-density lipoprotein receptor-related protein 1 at the blood-cerebrospinal fluid barrier. J Neurochem 118, 407–415 (2011).

30. Storck SE, et al. Endothelial LRP1 transports amyloid-β(1-42) across the blood-brain barrier. J Clin Invest 126, 123–136 (2016).

31. Saido T, Leissring MA. Proteolytic degradation of amyloid β-protein. Cold Spring Harb Perspect Med 2, a006379 (2012).

32. Olesen OF. Proteolytic degradation of microtubule associated protein tau by thrombin. Biochem Biophys Res Commun 201, 716–721 (1994).

33. Arai T, Guo JP, McGeer PL. Proteolysis of non-phosphorylated and phosphorylated tau by thrombin. J Biol Chem 280, 5145–5153 (2005).

34. Kanehisa M, Goto S. KEGG: kyoto encyclopedia of genes and genomes. Nucleic Acids Res 28, 27–30 (2000).

35. Kanehisa M, Furumichi M, Sato Y, Matsuura Y, Ishiguro-Watanabe M. KEGG: biological systems database as a model of the real world. Nucleic Acids Res 53, D672–D677 (2025).

36. Caccamo A, et al. mTOR regulates tau phosphorylation and degradation: implications for Alzheimer’s disease and other tauopathies. Aging Cell 12, 370– 380 (2013).

37. Ferrer I, et al. Phosphorylated map kinase (ERK1, ERK2) expression is associated with early tau deposition in neurones and glial cells, but not with increased nuclear DNA vulnerability and cell death, in Alzheimer disease, Pick’s disease, progressive supranuclear palsy and corticobasal degeneration. Brain Pathol 11, 144–158 (2001).

38. Reynolds CH, Betts JC, Blackstock WP, Nebreda AR, Anderton BH. Phosphorylation sites on tau identified by nanoelectrospray mass spectrometry: differences in vitro between the mitogen-activated protein kinases ERK2, c-Jun N-terminal kinase and P38, and glycogen synthase kinase-3beta. J Neurochem 74, 1587–1595 (2000).

39. Folstein MF, Folstein SE, McHugh PR. Mini-mental state: a practical method for grading the cognitive state of patients for the clinician. J Psychiat Res 12, 189– 198 (1975).

40. Liu H, et al. Acute sleep loss decreases CSF-to-blood clearance of Alzheimer’s disease biomarkers. Alzheimers Dement 19, 3055–3064 (2023).

41. Eastwood SM, et al. PrecivityAD2™ Blood Test: Analytical Validation of an LC-MS/MS Assay for Quantifying Plasma Phospho-tau217 and Non-Phospho-tau217 Peptide Concentrations That Are Used with Plasma Amyloid-β42/40 in a Multianalyte Assay with Algorithmic Analysis for Detecting Brain Amyloid Pathology. Diagnostics (Basel) 14, 1739 (2024).

42. Meyer MR, et al. Clinical validation of the PrecivityAD2 blood test: A mass spectrometry-based test with algorithm combining %p-tau217 and Aβ42/40 ratio to identify presence of brain amyloid. Alzheimers Dement 20, 3179–3192 (2024).

43. Kirmess KM, et al. The PrecivityAD™ test: Accurate and reliable LC-MS/MS assays for quantifying plasma amyloid beta 40 and 42 and apolipoprotein E proteotype for the assessment of brain amyloidosis. Clin Chim Acta 519, 267– 275 (2021).

44. Candia J, et al. Variability of 7K and 11K SomaScan Plasma Proteomics Assays. J Proteome Res 23, 5531–5539 (2024).

45. Feng W, et al. NULISA: a proteomic liquid biopsy platform with attomolar sensitivity and high multiplexing. Nat Commun 14, 7238 (2023).

46. Ibanez L, et al. Benchmarking of a multi-biomarker low-volume panel for Alzheimer’s disease and related dementia research. Alzheimers Dement 21, e14413 (2025).

