## Supplementary Figures and Tables for "Suvorexant Inversely Affects Phosphorylated Tau in Human Cerebrospinal Fluid and Plasma"

**SUPPLEMENTARY MATERIALS**

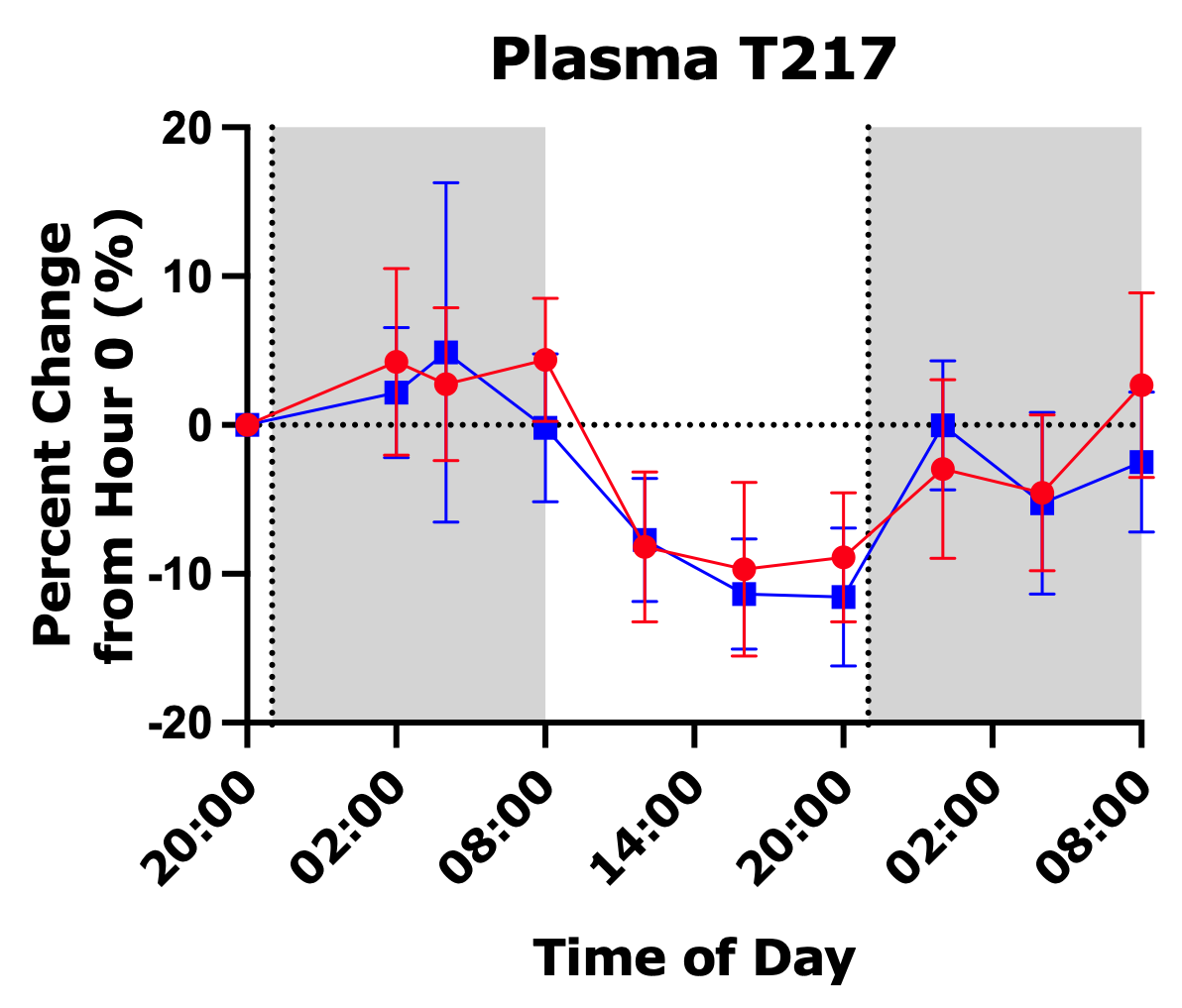

**Supplementary Figure 1. Plasma unphosphorylated tau-217 (T217) changes in cognitively unimpaired participants treated with suvorexant 20 mg** (blue squares, N = 12) **compared to placebo** (red circles, N = 12) over two consecutive nights**.** Percent change from hour 0 (20:00, day 1) in plasma T217. Points are group mean ± SEM. Shaded regions indicate the scheduled dark period (20:00–08:00); vertical dotted lines indicate administration of placebo or suvorexant at hour 1 (21:00, day 1) and hour 25 (21:00, day 2).

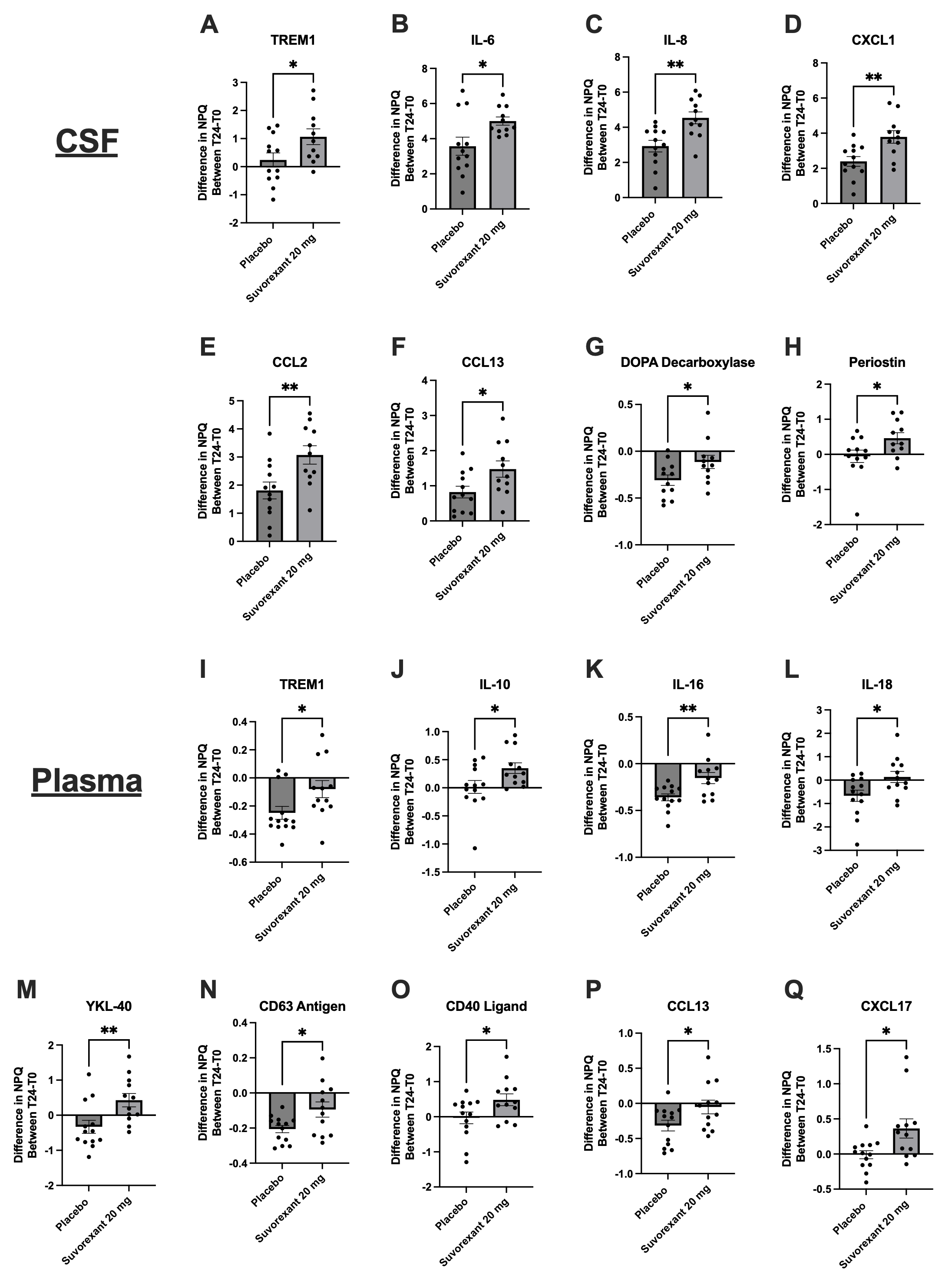

**Supplementary Figure 2. NULISA-measured inflammatory and immune proteins differing between suvorexant and placebo in CSF and plasma at hour 24.** Proteins measured on the NULISA CNS Disease Panel 120 that differed between treatment groups. Each panel shows the within-participant change in NULISA Protein Quantification (NPQ) units from hour 0 to hour 24 for placebo and suvorexant 20 mg. In the placebo group, there were 12 participants with CSF and 13 participants with plasma available for analysis. In the suvorexant group, there were 11 participants with CSF and 12 participants with plasma. Bars are group mean ± SEM with individual participants overlaid. (**A–H**) CSF: TREM1 (A), IL-6 (B), IL-8 (C), CXCL1 (D), CCL2 (E), CCL13 (F), DOPA decarboxylase (G), and periostin (H). (**I–Q**) Plasma: TREM1 (I), IL-10 (J), IL-16 (K), IL-18 (L), YKL-40 (M), CD63 antigen (N), CD40 ligand (O), CCL13 (P), and CXCL17 (Q). *p < 0.05, **p < 0.01. NPQ is a log₂-scale relative quantification, so differences are approximately log₂ fold changes. All comparisons are based on unadjusted p-values from t-tests; no protein survived correction for multiple comparisons. Note that y-axis scales differ between panels.

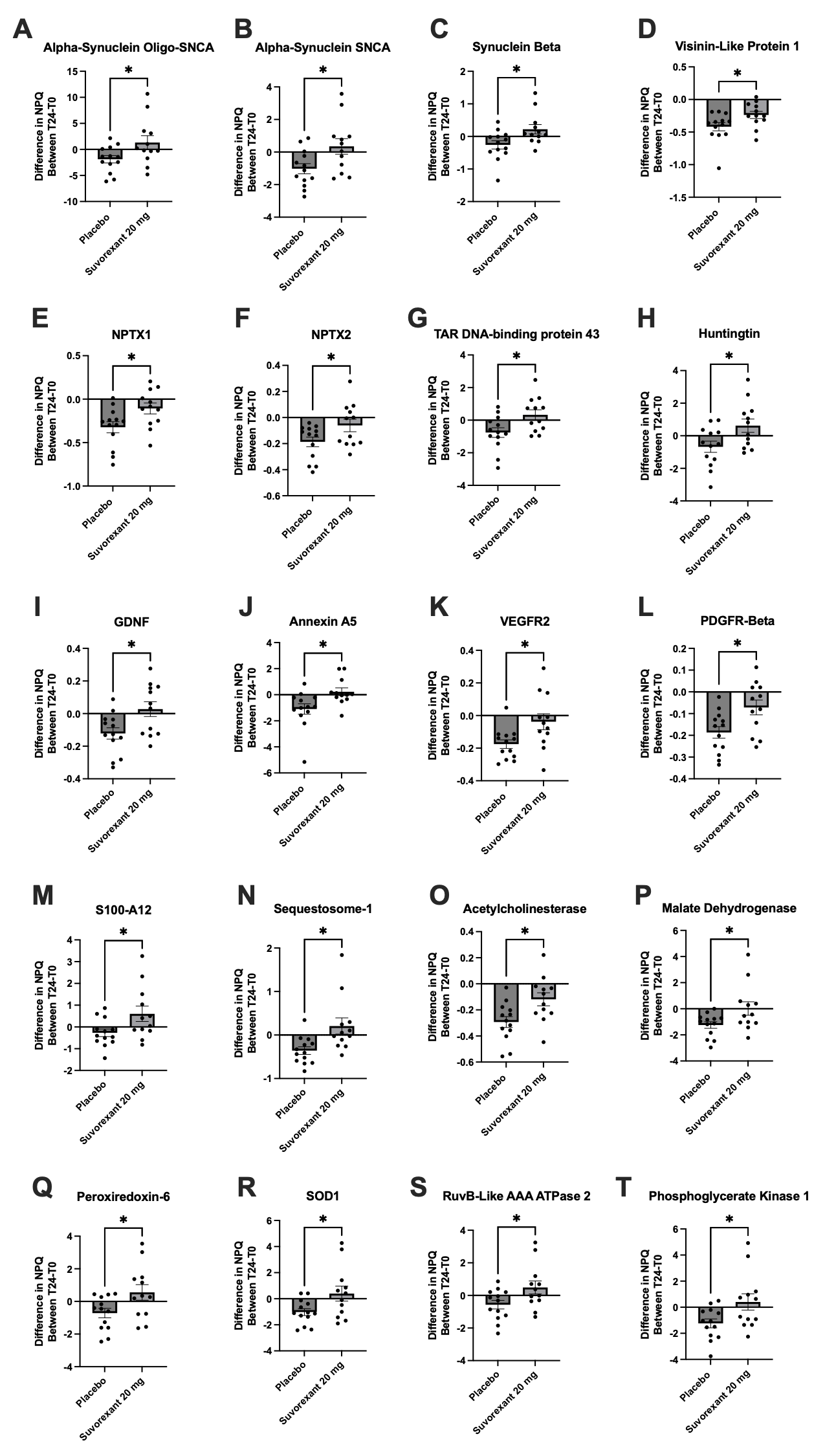

**Supplementary Figure 3. NULISA-measured neuronal, vascular, and metabolic plasma proteins differing between suvorexant and placebo at hour 24.** Additional proteins from the NULISA CNS Disease Panel 120 measured in plasma. Each panel shows the within-participant change in NPQ from hour 0 to hour 24 for placebo (N = 13) and suvorexant 20 mg (N = 12); bars are group mean ± SEM with individual participants overlaid. (**A**) oligomeric α-synuclein (oligo-SNCA), (**B**) total α-synuclein (SNCA), (**C**) β-synuclein, (**D**) visinin-like protein 1, (**E**) NPTX1, (**F**) NPTX2, (**G**) TAR DNA-binding protein 43, (**H**) huntingtin, (**I**) GDNF, (**J**) annexin A5, (**K**) VEGFR2, (**L**) PDGFR-β, (**M**) S100A12, (**N**) sequestosome-1, (**O**) acetylcholinesterase, (**P**) malate dehydrogenase, (**Q**) peroxiredoxin-6, (**R**) SOD1, (**S**) RuvB-like AAA ATPase 2, and (**T**) phosphoglycerate kinase 1. *p < 0.05. All comparisons based on unadjusted p-values from t-tests; no protein survived correction for multiple comparisons. Note that y-axis scales differ between panels.

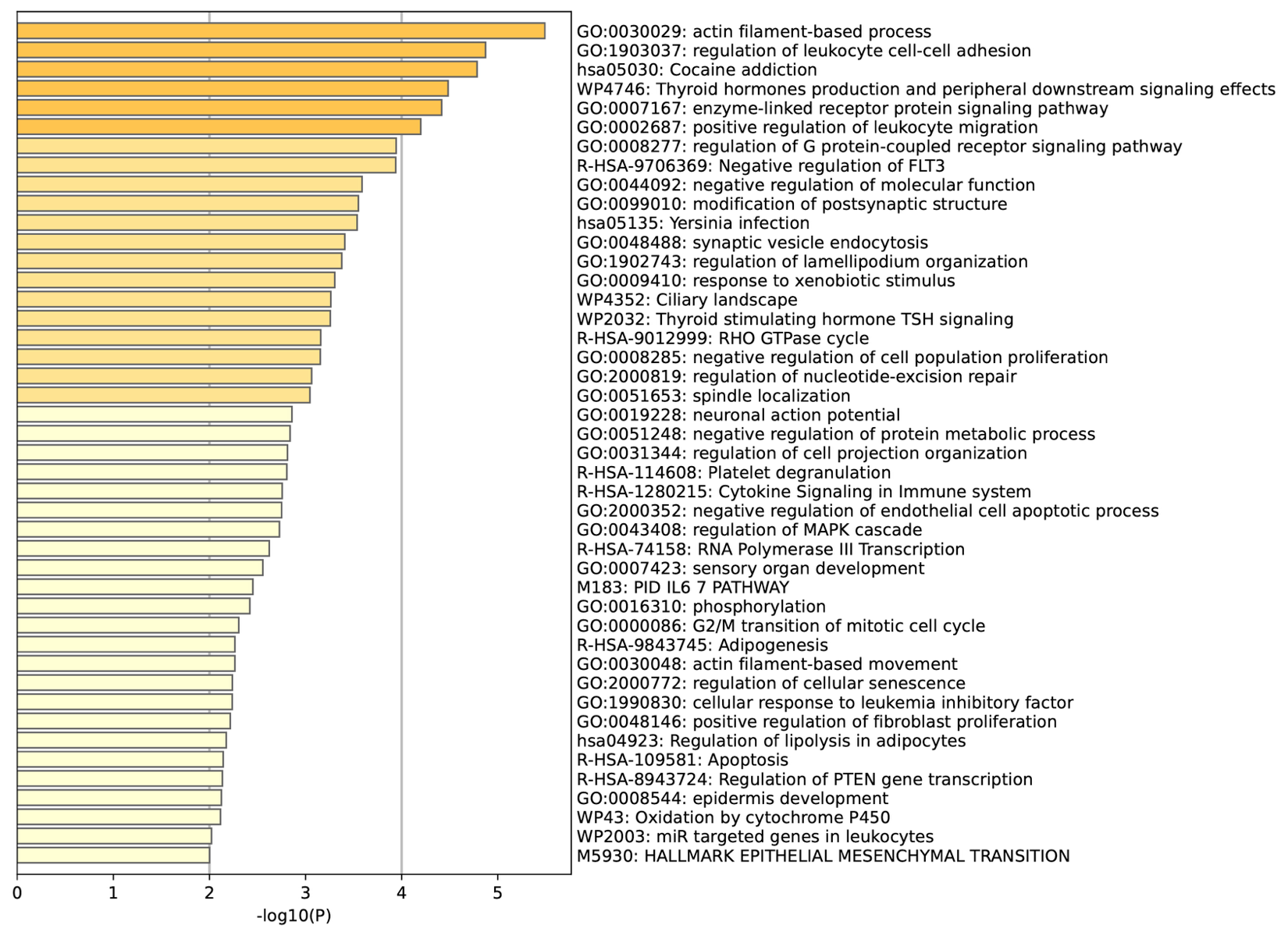

**Supplementary Figure 4. Complete Metascape enrichment results for CSF proteins increased by suvorexant 20 mg at hour 16.** All ontology terms enriched among CSF proteins higher in the suvorexant 20 mg group than in the placebo group at hour 16 relative to hour 0, ranked by −log₁₀(P) from the Metascape and shaded by significance. Terms required a minimum of three input proteins, an enrichment factor >1.5, and p < 0.05; the display is capped at the 100 most significant terms, and fewer than 100 terms met criteria for this contrast. Unlike the summary bar chart in Figure 4A, which shows one representative term per cluster, this figure shows all qualifying terms including redundant members of the same cluster. Input proteins were those with nominal p < 0.05 in the SomaScan limma contrast.

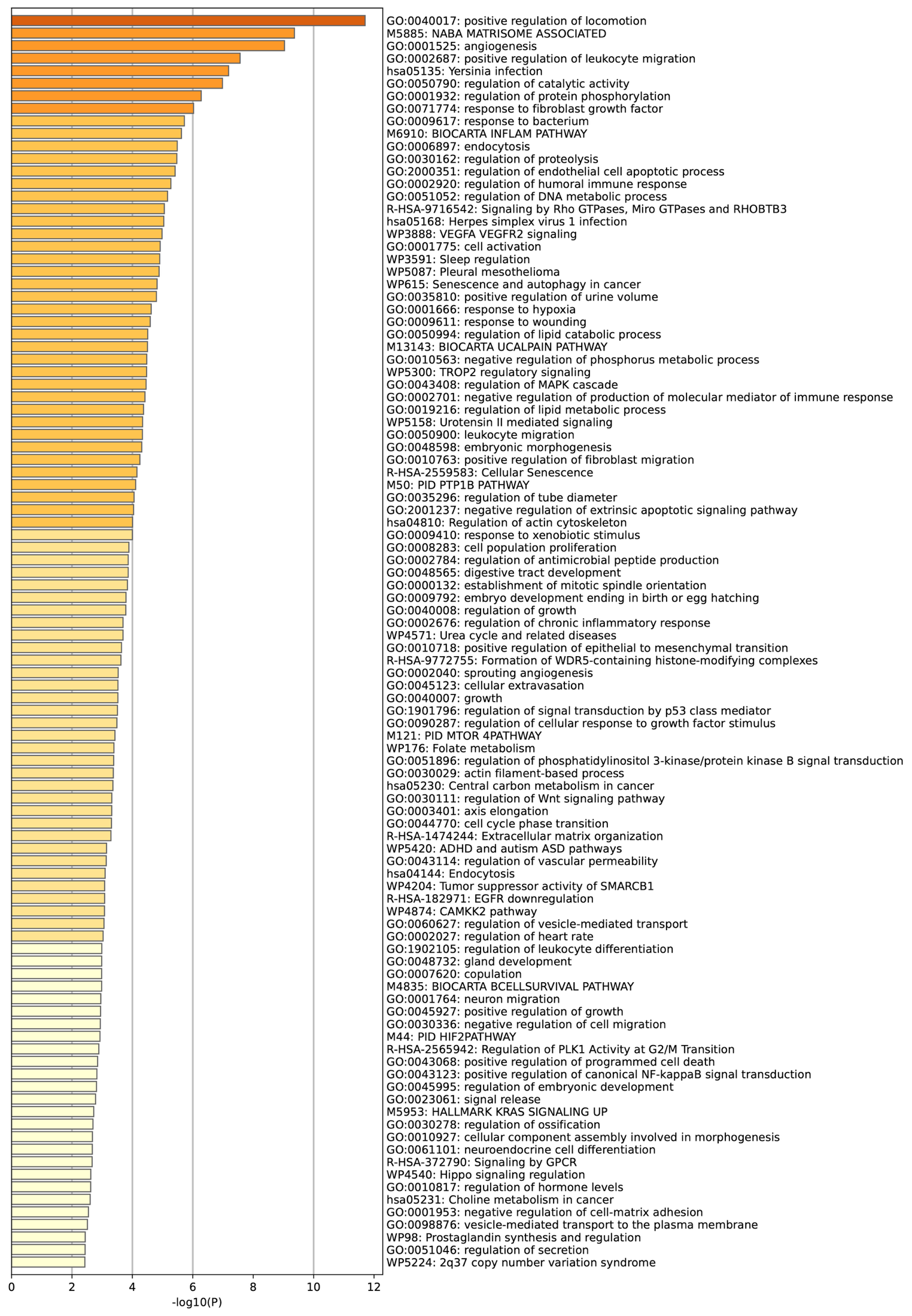

**Supplementary Figure 5. Complete Metascape enrichment results for CSF proteins increased by suvorexant 20 mg at hour 24.** All ontology terms enriched among CSF proteins higher in the suvorexant 20 mg group than in the placebo group at hour 24 relative to hour 0, ranked by −log₁₀(P) and shaded by significance, displaying the 100 most significant terms. Terms required a minimum of three input proteins, an enrichment factor >1.5, and p < 0.05; the display is capped at the 100 most significant terms, and fewer than 100 terms met criteria for this contrast. Input proteins were those with nominal p < 0.05 in the SomaScan limma contrast.

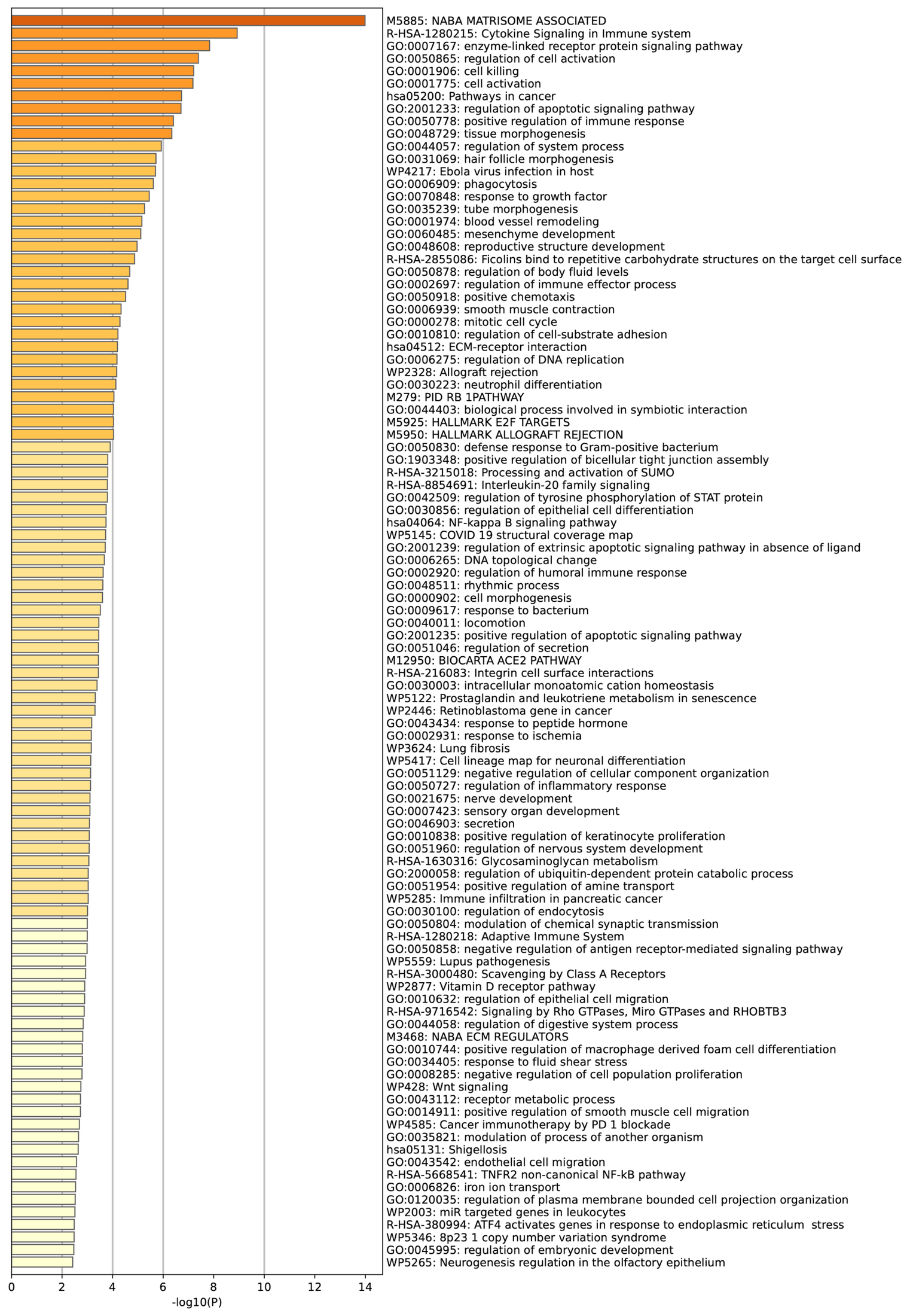

**Supplementary Figure 6. Complete Metascape enrichment results for CSF proteins decreased by suvorexant 20 mg at hour 24.** All ontology terms enriched among CSF proteins lower in the suvorexant 20 mg group than in the placebo group at hour 24 relative to hour 0, ranked by −log₁₀(P) and shaded by significance, displaying the 100 most significant terms. Terms required a minimum of three input proteins, an enrichment factor >1.5, and p < 0.05; the display is capped at the 100 most significant terms, and fewer than 100 terms met criteria for this contrast. Input proteins were those with nominal p < 0.05 in the SomaScan limma contrast.

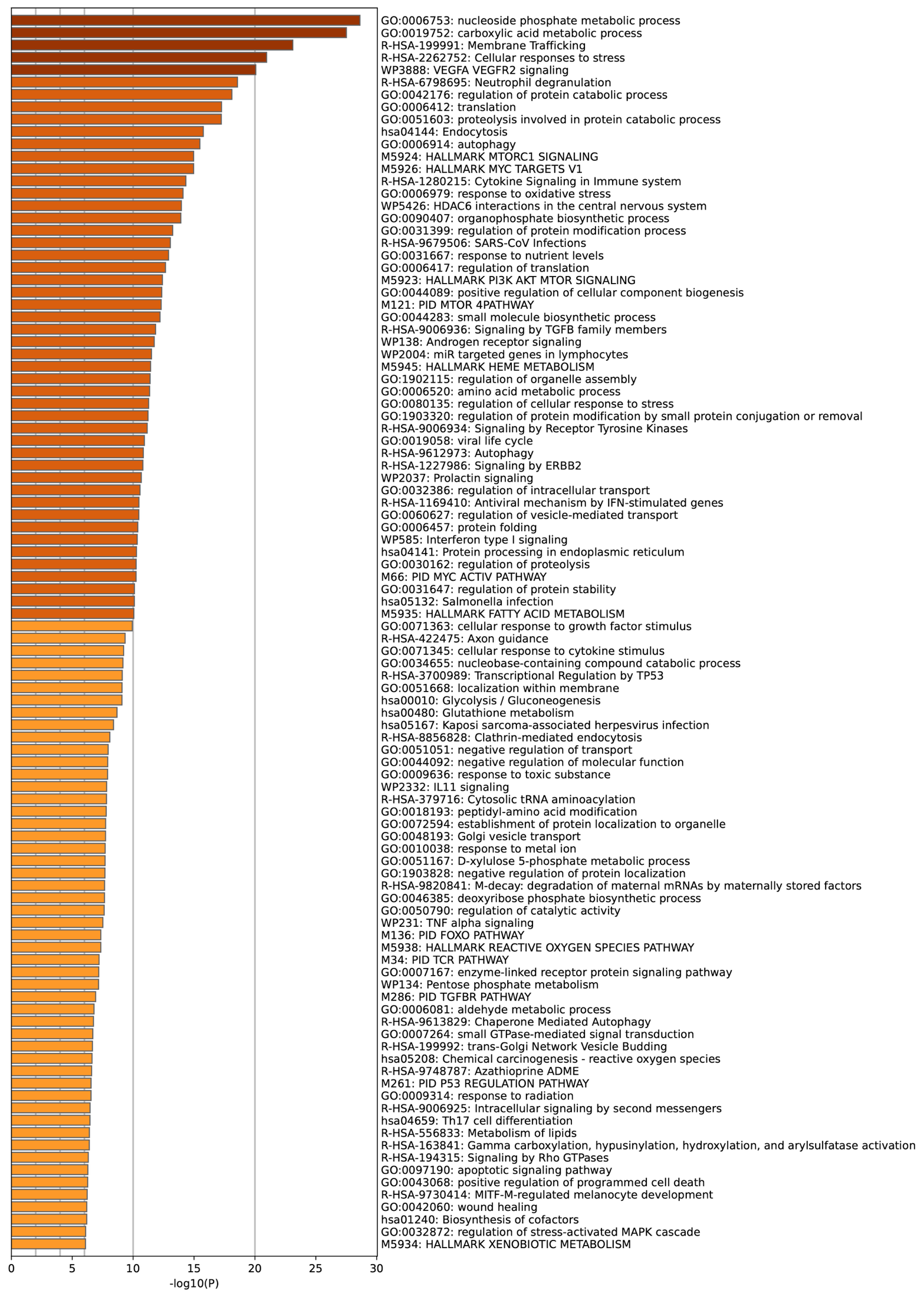

**Supplementary Figure 7. Complete Metascape enrichment results for plasma proteins increased by suvorexant 20 mg at hour 24.** All ontology terms enriched among plasma proteins higher in the suvorexant 20 mg group than in the placebo group at hour 24 relative to hour 0, ranked by −log₁₀(P) and shaded by significance, displaying the 100 most significant terms. Terms required a minimum of three input proteins, an enrichment factor >1.5, and p < 0.05; the display is capped at the 100 most significant terms, and fewer than 100 terms met criteria for this contrast. Input proteins were those with nominal p < 0.05 in the SomaScan limma contrast.

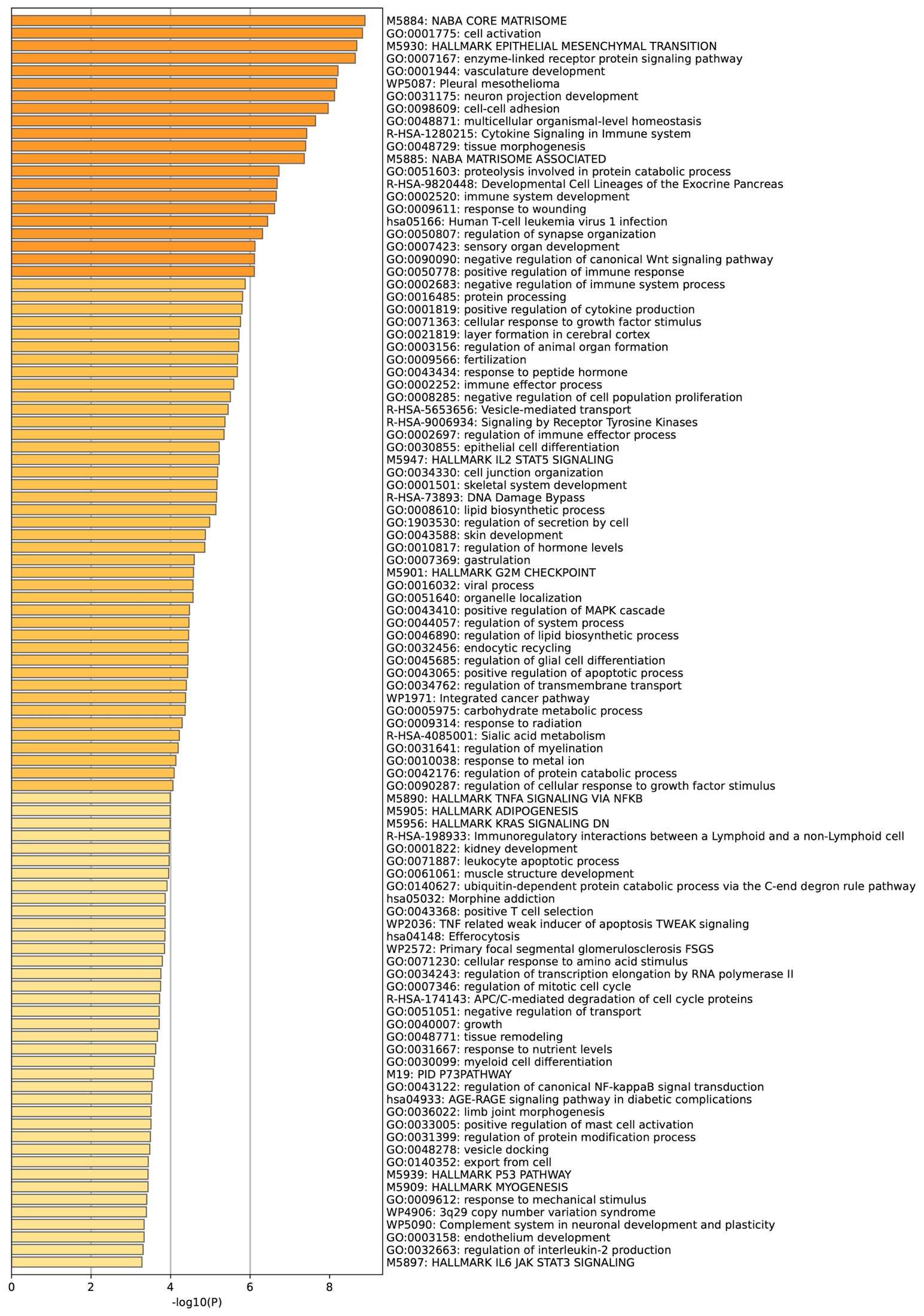

**Supplementary Figure 8. Complete Metascape enrichment results for plasma proteins decreased by suvorexant 20 mg at hour 24.** All ontology terms enriched among plasma proteins lower in the suvorexant 20 mg group than in the placebo group at hour 24 relative to hour 0, ranked by −log₁₀(P) and shaded by significance, displaying the 100 most significant terms. Terms required a minimum of three input proteins, an enrichment factor >1.5, and p < 0.05; the display is capped at the 100 most significant terms, and fewer than 100 terms met criteria for this contrast. Input proteins were those with nominal p < 0.05 in the SomaScan limma contrast.

**SUPPLEMENTARY TABLE 1**

| **CSF: Suvorexant 20 mg vs Placebo — Pathways with FDR < 0.05** | | | | |
| --- | --- | --- | --- | --- |
| *Sections color-coded by timepoint and analysis type:*  *• Change from hour 16 to hour 0 between suvorexant 20 mg and placebo: T16 = blue*  *• Change from hour 24 to hour 0 between suvorexant 20 mg and placebo: T24 = purple*  *• Sorted by False Discovery Rate (FDR) within each section* | | | | |
| **Pathway / Term** | **Set Size** | **Mean logFC vs Background** | **P-value** | **FDR** |
| ***T16 · GO Biological Process (8 pathways)*** | | | | |
| GO:0006412 Translation | 374 | 4.803 | 9.66e-7 | 0.0109 |
| GO:0006996 Organelle organization | 1915 | 4.366 | 3.67e-6 | 0.0246 |
| GO:0044085 Cellular component biogenesis | 1867 | 4.316 | 4.62e-6 | 0.0246 |
| GO:0007010 Cytoskeleton organization | 839 | 4.308 | 5.49e-6 | 0.0246 |
| GO:0022607 Cellular component assembly | 1749 | 4.117 | 2.31e-5 | 0.0412 |
| GO:0030029 Actin filament-based process | 496 | 4.088 | 2.44e-5 | 0.0412 |
| GO:0006139 Nucleobase-containing compound metabolic process | 2825 | 4.053 | 2.94e-5 | 0.0412 |
| GO:0016071 mRNA metabolic process | 392 | 4.007 | 4.02e-5 | 0.0477 |
| ***T16 · GO Molecular Function (24 pathways)*** | | | | |
| GO:0003723 RNA binding | 901 | 7.393 | 1.13e-13 | 3.11e-10 |
| GO:0000166 Nucleotide binding | 1207 | 5.820 | 5.15e-9 | 3.35e-6 |
| GO:1901265 Nucleoside phosphate binding | 1222 | 5.805 | 5.15e-9 | 3.35e-6 |
| GO:0017076 Purine nucleotide binding | 1112 | 5.649 | 1.18e-8 | 5.76e-6 |
| GO:1901363 Heterocyclic compound binding | 1308 | 5.622 | 1.18e-8 | 5.76e-6 |
| GO:0003676 Nucleic acid binding | 1840 | 5.341 | 6.95e-8 | 2.26e-5 |
| GO:0043168 Anion binding | 1405 | 5.240 | 1.32e-7 | 3.40e-5 |
| GO:0032555 Purine ribonucleotide binding | 1041 | 5.208 | 1.40e-7 | 3.62e-5 |
| GO:0035639 Purine ribonucleoside triphosphate binding | 1013 | 5.168 | 1.54e-7 | 3.99e-5 |
| GO:0032553 Ribonucleotide binding | 1052 | 5.140 | 1.61e-7 | 4.15e-5 |
| GO:0030554 Adenyl nucleotide binding | 895 | 4.728 | 1.19e-6 | 3.07e-4 |
| GO:0097367 Carbohydrate derivative binding | 1364 | 4.354 | 6.17e-6 | 0.0016 |
| GO:0003729 mRNA binding | 207 | 4.268 | 9.92e-6 | 0.0026 |
| GO:0032559 Adenyl ribonucleotide binding | 825 | 4.213 | 9.92e-6 | 0.0026 |
| GO:0005524 ATP binding | 798 | 4.202 | 9.92e-6 | 0.0026 |
| GO:0045296 Cadherin binding | 245 | 4.177 | 1.18e-5 | 0.0030 |
| GO:0035091 Phosphatidylinositol binding | 157 | 3.715 | 7.63e-5 | 0.0197 |
| GO:0019001 Guanyl nucleotide binding | 252 | 3.634 | 8.66e-5 | 0.0224 |
| GO:0032561 Guanyl ribonucleotide binding | 252 | 3.634 | 8.66e-5 | 0.0224 |
| GO:0008092 Cytoskeletal protein binding | 571 | 3.576 | 9.71e-5 | 0.0251 |
| GO:0005525 GTP binding | 240 | 3.517 | 1.22e-4 | 0.0315 |
| GO:0016874 Ligase activity | 94 | 3.502 | 1.45e-4 | 0.0374 |
| GO:0045182 Translation regulator activity | 84 | 3.505 | 1.45e-4 | 0.0374 |
| GO:0003925 G protein activity | 41 | 3.498 | 1.84e-4 | 0.0474 |
| ***T16 · KEGG Signaling & Metabolism (1 pathway)*** | | | | |
| hsa04144 Endocytosis | 184 | 3.892 | 5.91e-5 | 0.0149 |
| ***T24 · GO Molecular Function (1 pathway)*** | | | | |
| GO:0003723 RNA binding | 901 | 4.830 | 7.41e-7 | 0.0020 |

**SUPPLEMENTARY TABLE 2**

| **Plasma: Suvorexant 20 mg vs Placebo — Pathways with FDR < 0.05** | | | | |
| --- | --- | --- | --- | --- |
| *Sections color-coded by timepoint and analysis type:*   - *Change from hour 16 to hour 0 between suvorexant 20 mg and placebo: T16 = blue* - *Change from hour 24 to hour 0 between suvorexant 20 mg and placebo: T24 = purple* - *Sorted by False Discovery Rate (FDR) within each section* | | | | |
| **Pathway / Term** | **Set Size** | **Mean logFC vs Background** | **P-value** | **FDR** |
| ***T16 · GO Biological Process (12 pathways)*** | | | | |
| GO:0006082 organic acid metabolic process | 615 | 6.294 | 2.29e-10 | 1.08e-6 |
| GO:0043436 oxoacid metabolic process | 609 | 6.276 | 2.57e-10 | 1.08e-6 |
| GO:0019752 carboxylic acid metabolic process | 592 | 6.259 | 2.89e-10 | 1.08e-6 |
| GO:0044282 small molecule catabolic process | 269 | 5.217 | 1.42e-7 | 4.01e-4 |
| GO:0044281 small molecule metabolic process | 1169 | 5.080 | 2.06e-7 | 4.63e-4 |
| GO:0016054 organic acid catabolic process | 172 | 4.764 | 1.60e-6 | 2.58e-3 |
| GO:0046395 carboxylic acid catabolic process | 172 | 4.764 | 1.60e-6 | 2.58e-3 |
| GO:1901605 alpha-amino acid metabolic process | 152 | 4.622 | 3.25e-6 | 4.57e-3 |
| GO:0032787 monocarboxylic acid metabolic process | 410 | 4.508 | 3.84e-6 | 4.81e-3 |
| GO:0170033 L-amino acid metabolic process | 131 | 4.379 | 9.93e-6 | 1.12e-2 |
| GO:0170039 proteinogenic amino acid metabolic process | 126 | 4.321 | 1.29e-5 | 1.32e-2 |
| GO:0006520 amino acid metabolic process | 206 | 4.235 | 1.51e-5 | 1.42e-2 |
| ***T16 · GO Molecular Function (1 pathway)*** | | | | |
| GO:0016491 oxidoreductase activity | 451 | 4.292 | 9.92e-6 | 2.73e-2 |
| ***T16 · KEGG (9 pathways)*** | | | | |
| hsa01100 Metabolic pathways | 998 | 5.430 | 3.21e-8 | 8.10e-6 |
| hsa01200 Carbon metabolism | 98 | 3.781 | 1.13e-4 | 1.30e-2 |
| hsa00620 Pyruvate metabolism | 43 | 3.833 | 1.55e-4 | 1.30e-2 |
| hsa00071 Fatty acid degradation | 29 | 3.756 | 2.87e-4 | 1.81e-2 |
| hsa04144 Endocytosis | 184 | -3.693 | 1.28e-4 | 3.22e-2 |
| hsa00280 Valine, leucine and isoleucine degradation | 33 | 3.382 | 7.36e-4 | 3.71e-2 |
| hsa04146 Peroxisome | 51 | 3.103 | 1.34e-3 | 4.90e-2 |
| hsa00260 Glycine, serine and threonine metabolism | 31 | 3.116 | 1.53e-3 | 4.90e-2 |
| hsa00980 Metabolism of xenobiotics by cytochrome P450 | 56 | 3.037 | 1.56e-3 | 4.90e-2 |
| ***T24 · GO Biological Process (135 pathways)*** | | | | |
| GO:0006412 translation | 374 | 6.030 | 1.35e-9 | 1.52e-5 |
| GO:0055086 nucleobase-containing small molecule metabolic process | 458 | 5.731 | 7.04e-9 | 3.96e-5 |
| GO:0009056 catabolic process | 1641 | 5.534 | 1.69e-8 | 5.28e-5 |
| GO:0006753 nucleoside phosphate metabolic process | 438 | 5.559 | 1.87e-8 | 5.28e-5 |
| GO:0006996 organelle organization | 1915 | 5.377 | 4.03e-8 | 9.07e-5 |
| GO:0009117 nucleotide metabolic process | 346 | 5.363 | 5.88e-8 | 1.10e-4 |
| GO:0006796 phosphate-containing compound metabolic process | 1499 | 5.243 | 8.49e-8 | 1.21e-4 |
| GO:0006793 phosphorus metabolic process | 1500 | 5.241 | 8.57e-8 | 1.21e-4 |
| GO:0019538 protein metabolic process | 2908 | 5.123 | 1.56e-7 | 1.95e-4 |
| GO:0016236 macroautophagy | 222 | 5.095 | 2.73e-7 | 2.87e-4 |
| GO:0010608 post-transcriptional regulation of gene expression | 312 | 5.061 | 2.80e-7 | 2.87e-4 |
| GO:0046907 intracellular transport | 767 | 4.862 | 6.46e-7 | 5.68e-4 |
| GO:0019637 organophosphate metabolic process | 665 | 4.863 | 6.55e-7 | 5.68e-4 |
| GO:0032984 protein-containing complex disassembly | 158 | 4.912 | 7.92e-7 | 6.37e-4 |
| GO:0072521 purine-containing compound metabolic process | 376 | 4.812 | 9.24e-7 | 6.50e-4 |
| GO:0045184 establishment of protein localization | 995 | 4.777 | 9.61e-7 | 6.50e-4 |
| GO:0007010 cytoskeleton organization | 839 | 4.775 | 9.82e-7 | 6.50e-4 |
| GO:0044281 small molecule metabolic process | 1169 | 4.735 | 1.16e-6 | 7.28e-4 |
| GO:0019941 modification-dependent protein catabolic process | 380 | 4.750 | 1.24e-6 | 7.34e-4 |
| GO:0043632 modification-dependent macromolecule catabolic process | 382 | 4.726 | 1.39e-6 | 7.61e-4 |
| GO:0043933 protein-containing complex organization | 1052 | 4.681 | 1.52e-6 | 7.61e-4 |
| GO:0006417 regulation of translation | 228 | 4.725 | 1.57e-6 | 7.61e-4 |
| GO:0008104 protein localization | 1489 | 4.652 | 1.72e-6 | 7.61e-4 |
| GO:0006914 autophagy | 354 | 4.682 | 1.73e-6 | 7.61e-4 |
| GO:0061919 process utilizing autophagic mechanism | 354 | 4.682 | 1.73e-6 | 7.61e-4 |
| GO:0051641 cellular localization | 1939 | 4.645 | 1.76e-6 | 7.61e-4 |
| GO:0070727 cellular macromolecule localization | 1497 | 4.618 | 2.02e-6 | 8.26e-4 |
| GO:0009057 macromolecule catabolic process | 861 | 4.622 | 2.05e-6 | 8.26e-4 |
| GO:0044248 cellular catabolic process | 732 | 4.615 | 2.15e-6 | 8.36e-4 |
| GO:0051649 establishment of localization in cell | 1091 | 4.590 | 2.35e-6 | 8.80e-4 |
| GO:0051603 proteolysis involved in protein catabolic process | 480 | 4.595 | 2.48e-6 | 9.00e-4 |
| GO:0071705 nitrogen compound transport | 1033 | 4.553 | 2.81e-6 | 9.69e-4 |
| GO:0006163 purine nucleotide metabolic process | 300 | 4.581 | 2.91e-6 | 9.69e-4 |
| GO:0036258 multivesicular body assembly | 26 | 5.259 | 2.93e-6 | 9.69e-4 |
| GO:0015031 protein transport | 845 | 4.521 | 3.31e-6 | 1.07e-3 |
| GO:0006511 ubiquitin-dependent protein catabolic process | 372 | 4.511 | 3.81e-6 | 1.17e-3 |
| GO:0006413 translational initiation | 78 | 4.667 | 3.86e-6 | 1.17e-3 |
| GO:0036257 multivesicular body organization | 27 | 5.132 | 4.01e-6 | 1.19e-3 |
| GO:0090407 organophosphate biosynthetic process | 348 | 4.459 | 4.89e-6 | 1.41e-3 |
| GO:0030163 protein catabolic process | 647 | 4.435 | 5.04e-6 | 1.41e-3 |
| GO:0016197 endosomal transport | 183 | 4.487 | 5.12e-6 | 1.41e-3 |
| GO:0002181 cytoplasmic translation | 92 | 4.505 | 6.33e-6 | 1.70e-3 |
| GO:0019076 viral release from host cell | 25 | 4.941 | 7.92e-6 | 2.03e-3 |
| GO:0035891 exit from host cell | 25 | 4.941 | 7.92e-6 | 2.03e-3 |
| GO:0070925 organelle assembly | 535 | 4.310 | 8.96e-6 | 2.24e-3 |
| GO:0007032 endosome organization | 65 | 4.494 | 9.61e-6 | 2.35e-3 |
| GO:0032509 endosome transport via multivesicular body sorting pathway | 30 | 4.802 | 1.00e-5 | 2.40e-3 |
| GO:0006457 protein folding | 175 | 4.287 | 1.21e-5 | 2.84e-3 |
| GO:0019693 ribose phosphate metabolic process | 253 | 4.235 | 1.40e-5 | 3.22e-3 |
| GO:0071985 multivesicular body sorting pathway | 34 | 4.623 | 1.44e-5 | 3.22e-3 |
| GO:0044087 regulation of cellular component biogenesis | 655 | 4.193 | 1.47e-5 | 3.22e-3 |
| GO:0022607 cellular component assembly | 1749 | 4.181 | 1.48e-5 | 3.22e-3 |
| GO:0044085 cellular component biogenesis | 1867 | 4.170 | 1.56e-5 | 3.31e-3 |
| GO:0033036 macromolecule localization | 1745 | 4.102 | 2.10e-5 | 4.38e-3 |
| GO:1902115 regulation of organelle assembly | 136 | 4.167 | 2.16e-5 | 4.42e-3 |
| GO:0051246 regulation of protein metabolic process | 1348 | 4.078 | 2.33e-5 | 4.63e-3 |
| GO:0006886 intracellular protein transport | 400 | 4.095 | 2.34e-5 | 4.63e-3 |
| GO:0051493 regulation of cytoskeleton organization | 321 | 4.094 | 2.41e-5 | 4.68e-3 |
| GO:0010256 endomembrane system organization | 343 | 4.072 | 2.64e-5 | 4.98e-3 |
| GO:0009894 regulation of catabolic process | 629 | 4.056 | 2.65e-5 | 4.98e-3 |
| GO:1901293 nucleoside phosphate biosynthetic process | 196 | 4.086 | 2.76e-5 | 5.09e-3 |
| GO:0016192 vesicle-mediated transport | 984 | 3.959 | 3.90e-5 | 7.08e-3 |
| GO:0033043 regulation of organelle organization | 692 | 3.949 | 4.14e-5 | 7.39e-3 |
| GO:0030036 actin cytoskeleton organization | 454 | 3.951 | 4.20e-5 | 7.39e-3 |
| GO:0032506 cytokinetic process | 36 | 4.183 | 5.41e-5 | 9.33e-3 |
| GO:0140694 non-membrane-bounded organelle assembly | 246 | 3.903 | 5.47e-5 | 9.33e-3 |
| GO:0007017 microtubule-based process | 487 | 3.832 | 6.81e-5 | 1.11e-2 |
| GO:0019068 virion assembly | 29 | 4.179 | 6.83e-5 | 1.11e-2 |
| GO:0009259 ribonucleotide metabolic process | 246 | 3.848 | 6.88e-5 | 1.11e-2 |
| GO:0006139 nucleobase-containing compound metabolic process | 2825 | 3.811 | 6.99e-5 | 1.11e-2 |
| GO:0065003 protein-containing complex assembly | 928 | 3.810 | 7.17e-5 | 1.11e-2 |
| GO:0032535 regulation of cellular component size | 230 | 3.835 | 7.21e-5 | 1.11e-2 |
| GO:0043254 regulation of protein-containing complex assembly | 270 | 3.831 | 7.22e-5 | 1.11e-2 |
| GO:0031468 nuclear membrane reassembly | 24 | 4.202 | 7.92e-5 | 1.21e-2 |
| GO:0045324 late endosome to vacuole transport | 26 | 4.194 | 8.33e-5 | 1.23e-2 |
| GO:0022411 cellular component disassembly | 289 | 3.792 | 8.36e-5 | 1.23e-2 |
| GO:0009123 nucleoside monophosphate metabolic process | 59 | 3.923 | 8.42e-5 | 1.23e-2 |
| GO:0007051 spindle organization | 125 | 3.814 | 9.04e-5 | 1.30e-2 |
| GO:0016050 vesicle organization | 236 | 3.765 | 9.53e-5 | 1.34e-2 |
| GO:0007015 actin filament organization | 282 | 3.757 | 9.54e-5 | 1.34e-2 |
| GO:0009167 purine ribonucleoside monophosphate metabolic process | 32 | 4.046 | 9.92e-5 | 1.37e-2 |
| GO:0008333 endosome to lysosome transport | 40 | 3.966 | 9.97e-5 | 1.37e-2 |
| GO:0009161 ribonucleoside monophosphate metabolic process | 47 | 3.905 | 1.05e-4 | 1.43e-2 |
| GO:0009150 purine ribonucleotide metabolic process | 231 | 3.732 | 1.09e-4 | 1.46e-2 |
| GO:0009165 nucleotide biosynthetic process | 140 | 3.752 | 1.11e-4 | 1.48e-2 |
| GO:0009126 purine nucleoside monophosphate metabolic process | 33 | 3.973 | 1.20e-4 | 1.57e-2 |
| GO:0032510 endosome to lysosome transport via multivesicular body sorting pathway | 15 | 4.390 | 1.21e-4 | 1.57e-2 |
| GO:0097212 lysosomal membrane organization | 12 | 4.522 | 1.31e-4 | 1.66e-2 |
| GO:0061136 regulation of proteasomal protein catabolic process | 134 | 3.707 | 1.32e-4 | 1.66e-2 |
| GO:0051469 vesicle fusion with vacuole | 11 | 4.598 | 1.38e-4 | 1.68e-2 |
| GO:0061763 multivesicular body-lysosome fusion | 11 | 4.598 | 1.38e-4 | 1.68e-2 |
| GO:0097576 vacuole fusion | 11 | 4.598 | 1.38e-4 | 1.68e-2 |
| GO:1902410 mitotic cytokinetic process | 24 | 4.025 | 1.39e-4 | 1.68e-2 |
| GO:1903050 regulation of proteolysis involved in protein catabolic process | 157 | 3.679 | 1.40e-4 | 1.68e-2 |
| GO:0030029 actin filament-based process | 496 | 3.638 | 1.45e-4 | 1.72e-2 |
| GO:0006520 amino acid metabolic process | 206 | 3.621 | 1.67e-4 | 1.95e-2 |
| GO:0000281 mitotic cytokinesis | 57 | 3.727 | 1.68e-4 | 1.95e-2 |
| GO:0061077 chaperone-mediated protein folding | 65 | 3.698 | 1.73e-4 | 1.99e-2 |
| GO:0046755 viral budding | 20 | 4.027 | 1.84e-4 | 2.09e-2 |
| GO:0061952 midbody abscission | 17 | 4.097 | 1.91e-4 | 2.15e-2 |
| GO:0061024 membrane organization | 497 | 3.556 | 1.98e-4 | 2.20e-2 |
| GO:0090148 membrane fission | 30 | 3.837 | 2.03e-4 | 2.24e-2 |
| GO:0006446 regulation of translational initiation | 54 | 3.671 | 2.06e-4 | 2.24e-2 |
| GO:0039702 viral budding via host ESCRT complex | 17 | 4.086 | 2.07e-4 | 2.24e-2 |
| GO:0051258 protein polymerization | 182 | 3.555 | 2.17e-4 | 2.32e-2 |
| GO:0046390 ribose phosphate biosynthetic process | 101 | 3.591 | 2.19e-4 | 2.32e-2 |
| GO:0032880 regulation of protein localization | 591 | 3.525 | 2.20e-4 | 2.32e-2 |
| GO:0000226 microtubule cytoskeleton organization | 332 | 3.524 | 2.29e-4 | 2.39e-2 |
| GO:0060341 regulation of cellular localization | 644 | 3.499 | 2.42e-4 | 2.50e-2 |
| GO:0019752 carboxylic acid metabolic process | 592 | 3.481 | 2.59e-4 | 2.64e-2 |
| GO:0042176 regulation of protein catabolic process | 250 | 3.495 | 2.60e-4 | 2.64e-2 |
| GO:0009408 response to heat | 77 | 3.535 | 2.82e-4 | 2.83e-2 |
| GO:0010638 positive regulation of organelle organization | 318 | 3.450 | 3.01e-4 | 3.00e-2 |
| GO:0060627 regulation of vesicle-mediated transport | 379 | 3.442 | 3.05e-4 | 3.02e-2 |
| GO:0051247 positive regulation of protein metabolic process | 752 | 3.431 | 3.09e-4 | 3.03e-2 |
| GO:0010498 proteasomal protein catabolic process | 342 | 3.428 | 3.23e-4 | 3.14e-2 |
| GO:0043162 ubiquitin-dependent protein catabolic process via the multivesicular body sorting pathway | 28 | 3.650 | 3.70e-4 | 3.53e-2 |
| GO:0046761 viral budding from plasma membrane | 14 | 3.968 | 3.72e-4 | 3.53e-2 |
| GO:0033365 protein localization to organelle | 553 | 3.382 | 3.73e-4 | 3.53e-2 |
| GO:0043436 oxoacid metabolic process | 609 | 3.378 | 3.78e-4 | 3.54e-2 |
| GO:1902903 regulation of supramolecular fiber organization | 238 | 3.383 | 3.92e-4 | 3.65e-2 |
| GO:0061640 cytoskeleton-dependent cytokinesis | 78 | 3.435 | 3.96e-4 | 3.66e-2 |
| GO:0071763 nuclear membrane organization | 36 | 3.542 | 4.14e-4 | 3.79e-2 |
| GO:0007052 mitotic spindle organization | 92 | 3.409 | 4.19e-4 | 3.81e-2 |
| GO:1901673 regulation of mitotic spindle assembly | 21 | 3.682 | 4.28e-4 | 3.86e-2 |
| GO:0006082 organic acid metabolic process | 615 | 3.307 | 4.85e-4 | 4.34e-2 |
| GO:0097352 autophagosome maturation | 39 | 3.464 | 4.92e-4 | 4.37e-2 |
| GO:0072522 purine-containing compound biosynthetic process | 151 | 3.331 | 4.98e-4 | 4.38e-2 |
| GO:0002183 cytoplasmic translational initiation | 28 | 3.513 | 5.10e-4 | 4.45e-2 |
| GO:0090307 mitotic spindle assembly | 51 | 3.404 | 5.14e-4 | 4.45e-2 |
| GO:0031334 positive regulation of protein-containing complex assembly | 135 | 3.316 | 5.30e-4 | 4.53e-2 |
| GO:0046753 non-lytic viral release | 15 | 3.778 | 5.31e-4 | 4.53e-2 |
| GO:0009168 purine ribonucleoside monophosphate biosynthetic process | 15 | 3.781 | 5.38e-4 | 4.55e-2 |
| GO:0036211 protein modification process | 1614 | 3.270 | 5.43e-4 | 4.56e-2 |
| GO:0034655 nucleobase-containing compound catabolic process | 323 | 3.268 | 5.71e-4 | 4.76e-2 |
| ***T24 · GO Molecular Function (55 pathways)*** | | | | |
| GO:0003723 RNA binding | 901 | 7.608 | 2.31e-14 | 6.36e-11 |
| GO:1901363 heterocyclic compound binding | 1308 | 7.152 | 5.66e-13 | 7.05e-10 |
| GO:1901265 nucleoside phosphate binding | 1222 | 7.112 | 7.68e-13 | 7.05e-10 |
| GO:0000166 nucleotide binding | 1207 | 7.038 | 1.31e-12 | 8.99e-10 |
| GO:0017076 purine nucleotide binding | 1112 | 6.820 | 6.01e-12 | 3.31e-9 |
| GO:0043168 anion binding | 1405 | 6.509 | 4.52e-11 | 1.56e-8 |
| GO:0032553 ribonucleotide binding | 1052 | 6.513 | 4.70e-11 | 1.56e-8 |
| GO:0035639 purine ribonucleoside triphosphate binding | 1013 | 6.513 | 4.74e-11 | 1.56e-8 |
| GO:0032555 purine ribonucleotide binding | 1041 | 6.501 | 5.09e-11 | 1.56e-8 |
| GO:0045296 cadherin binding | 245 | 6.494 | 1.19e-10 | 3.28e-8 |
| GO:0030554 adenyl nucleotide binding | 895 | 5.991 | 1.28e-9 | 3.22e-7 |
| GO:0005524 ATP binding | 798 | 5.748 | 5.50e-9 | 1.26e-6 |
| GO:0038023 signaling receptor activity | 631 | -5.987 | 1.53e-9 | 1.49e-6 |
| GO:0060089 molecular transducer activity | 631 | -5.987 | 1.53e-9 | 1.49e-6 |
| GO:0004888 transmembrane signaling receptor activity | 457 | -6.006 | 1.62e-9 | 1.49e-6 |
| GO:0032559 adenyl ribonucleotide binding | 825 | 5.661 | 9.02e-9 | 1.91e-6 |
| GO:0045182 translation regulator activity | 84 | 5.667 | 4.24e-8 | 8.34e-6 |
| GO:0090079 translation regulator activity, nucleic acid binding | 66 | 5.572 | 1.03e-7 | 1.90e-5 |
| GO:0008135 translation factor activity, RNA binding | 55 | 5.395 | 3.16e-7 | 5.43e-5 |
| GO:0097367 carbohydrate derivative binding | 1364 | 4.952 | 3.91e-7 | 6.33e-5 |
| GO:0019899 enzyme binding | 1279 | 4.913 | 4.77e-7 | 7.30e-5 |
| GO:0008092 cytoskeletal protein binding | 571 | 4.864 | 6.58e-7 | 9.54e-5 |
| GO:0050839 cell adhesion molecule binding | 439 | 4.735 | 1.29e-6 | 1.78e-4 |
| GO:0030545 signaling receptor regulator activity | 506 | -4.934 | 4.92e-7 | 2.83e-4 |
| GO:0048018 receptor ligand activity | 469 | -4.930 | 5.14e-7 | 2.83e-4 |
| GO:0042802 identical protein binding | 1535 | 4.553 | 2.75e-6 | 3.60e-4 |
| GO:0003743 translation initiation factor activity | 34 | 5.030 | 3.21e-6 | 4.02e-4 |
| GO:0030546 signaling receptor activator activity | 474 | -4.691 | 1.62e-6 | 7.42e-4 |
| GO:0044389 ubiquitin-like protein ligase binding | 232 | 4.412 | 6.58e-6 | 7.88e-4 |
| GO:0031625 ubiquitin protein ligase binding | 215 | 4.355 | 8.62e-6 | 9.89e-4 |
| GO:0003676 nucleic acid binding | 1840 | 4.260 | 1.05e-5 | 1.15e-3 |
| GO:0035091 phosphatidylinositol binding | 157 | 4.216 | 1.69e-5 | 1.79e-3 |
| GO:0016874 ligase activity | 94 | 4.237 | 2.03e-5 | 2.07e-3 |
| GO:0000287 magnesium ion binding | 153 | 4.088 | 2.87e-5 | 2.83e-3 |
| GO:0005543 phospholipid binding | 306 | 3.965 | 4.15e-5 | 3.94e-3 |
| GO:0003729 mRNA binding | 207 | 3.964 | 4.39e-5 | 4.03e-3 |
| GO:0019001 guanyl nucleotide binding | 252 | 3.900 | 5.55e-5 | 4.77e-3 |
| GO:0032561 guanyl ribonucleotide binding | 252 | 3.900 | 5.55e-5 | 4.77e-3 |
| GO:0019904 protein domain specific binding | 429 | 3.857 | 6.18e-5 | 5.16e-3 |
| GO:0005125 cytokine activity | 209 | -4.226 | 1.60e-5 | 6.30e-3 |
| GO:0005525 GTP binding | 240 | 3.748 | 1.02e-4 | 8.23e-3 |
| GO:0008289 lipid binding | 538 | 3.691 | 1.18e-4 | 8.80e-3 |
| GO:0016462 pyrophosphatase activity | 427 | 3.687 | 1.21e-4 | 8.80e-3 |
| GO:0016817 hydrolase activity, acting on acid anhydrides | 427 | 3.687 | 1.21e-4 | 8.80e-3 |
| GO:0016818 hydrolase activity, acting on acid anhydrides, in phosphorus-containing anhydrides | 427 | 3.687 | 1.21e-4 | 8.80e-3 |
| GO:0003925 G protein activity | 41 | 3.774 | 1.81e-4 | 1.28e-2 |
| GO:0016787 hydrolase activity | 1431 | 3.408 | 3.32e-4 | 2.29e-2 |
| GO:1901981 phosphatidylinositol phosphate binding | 102 | 3.444 | 3.55e-4 | 2.38e-2 |
| GO:0051219 phosphoprotein binding | 75 | 3.448 | 3.84e-4 | 2.52e-2 |
| GO:0017111 ribonucleoside triphosphate phosphatase activity | 385 | 3.342 | 4.37e-4 | 2.79e-2 |
| GO:0048156 tau protein binding | 25 | 3.573 | 4.46e-4 | 2.79e-2 |
| GO:0008236 serine-type peptidase activity | 143 | -3.736 | 1.15e-4 | 3.73e-2 |
| GO:0017171 serine hydrolase activity | 144 | -3.720 | 1.22e-4 | 3.73e-2 |
| GO:0003779 actin binding | 265 | 3.240 | 6.37e-4 | 3.90e-2 |
| GO:0005201 extracellular matrix structural constituent | 118 | -3.673 | 1.64e-4 | 4.51e-2 |
| ***T24 · KEGG (27 pathways)*** | | | | |
| hsa04144 Endocytosis | 184 | 5.647 | 1.86e-8 | 4.69e-6 |
| hsa01100 Metabolic pathways | 998 | 4.795 | 8.77e-7 | 1.10e-4 |
| hsa04060 Cytokine-cytokine receptor interaction | 262 | -4.739 | 1.56e-6 | 3.94e-4 |
| hsa01200 Carbon metabolism | 98 | 4.524 | 5.70e-6 | 4.79e-4 |
| hsa04974 Protein digestion and absorption | 60 | -4.320 | 1.76e-5 | 2.22e-3 |
| hsa00230 Purine metabolism | 91 | 3.840 | 9.23e-5 | 4.88e-3 |
| hsa01232 Nucleotide metabolism | 68 | 3.871 | 9.67e-5 | 4.88e-3 |
| hsa04722 Neurotrophin signaling pathway | 98 | 3.725 | 1.30e-4 | 5.48e-3 |
| hsa04071 Sphingolipid signaling pathway | 74 | 3.533 | 2.86e-4 | 1.01e-2 |
| hsa04910 Insulin signaling pathway | 92 | 3.478 | 3.21e-4 | 1.01e-2 |
| hsa00620 Pyruvate metabolism | 43 | 3.501 | 4.02e-4 | 1.12e-2 |
| hsa01230 Biosynthesis of amino acids | 64 | 3.370 | 5.19e-4 | 1.31e-2 |
| hsa04370 VEGF signaling pathway | 48 | 3.344 | 6.34e-4 | 1.45e-2 |
| hsa04062 Chemokine signaling pathway | 136 | 3.161 | 8.84e-4 | 1.86e-2 |
| hsa01240 Biosynthesis of cofactors | 115 | 3.101 | 1.10e-3 | 2.14e-2 |
| hsa04080 Neuroactive ligand-receptor interaction | 154 | -3.481 | 3.00e-4 | 2.52e-2 |
| hsa04012 ErbB signaling pathway | 76 | 2.993 | 1.63e-3 | 2.74e-2 |
| hsa04666 Fc gamma R-mediated phagocytosis | 68 | 3.004 | 1.63e-3 | 2.74e-2 |
| hsa04137 Mitophagy - animal | 72 | 2.913 | 2.13e-3 | 3.35e-2 |
| hsa04141 Protein processing in endoplasmic reticulum | 121 | 2.858 | 2.34e-3 | 3.47e-2 |
| hsa04662 B cell receptor signaling pathway | 67 | 2.855 | 2.57e-3 | 3.58e-2 |
| hsa00970 Aminoacyl-tRNA biosynthesis | 32 | 2.912 | 2.74e-3 | 3.58e-2 |
| hsa00480 Glutathione metabolism | 48 | 2.838 | 2.91e-3 | 3.58e-2 |
| hsa04140 Autophagy - animal | 104 | 2.762 | 3.14e-3 | 3.58e-2 |
| hsa04150 mTOR signaling pathway | 97 | 2.756 | 3.24e-3 | 3.58e-2 |
| hsa04664 Fc epsilon RI signaling pathway | 55 | 2.782 | 3.26e-3 | 3.58e-2 |
| hsa00630 Glyoxylate and dicarboxylate metabolism | 27 | 2.781 | 3.81e-3 | 4.00e-2 |
